# InfEHR: Resolving Clinical Uncertainty through Deep Geometric Learning on Electronic Health Records

**DOI:** 10.1101/2025.01.31.25321471

**Authors:** Justin Kauffman, Emma Holmes, Akhil Vaid, Alexander W Charney, Patricia Kovatch, Joshua Lampert, Ankit Sakhuja, Marinka Zitnik, Benjamin S Glicksberg, Ira Hofer, Girish N Nadkarni

**Affiliations:** The Windreich Department of Artificial Intelligence and Human Health, Icahn School of Medicine at Mount Sinai, New York, New York, USA; The Hasso Plattner Institute of Digital Health at Mount Sinai, Icahn School of Medicine at Mount Sinai, New York, New York, USA; The Charles Bronfman Institute for Personalized Medicine at Mount Sinai, Icahn School of Medicine at Mount Sinai, New York, New York, USA; Department of Data-Driven and Digital Medicine, Icahn School of Medicine at Mount Sinai, New York, New York, USA; Institute for Critical Care Medicine, Icahn School of Medicine at Mount Sinai, New York, New York, USA; Division of Newborn Medicine, Icahn School of Medicine at Mount Sinai, New York, New York, USA; Department of Medicine, Icahn School of Medicine at Mount Sinai, New York, New York, USA; Department of Surgery, Icahn School of Medicine at Mount Sinai, New York, New York, USA; Valentin Fuster Heart Hospital, Icahn School of Medicine at Mount Sinai, New York, New York, USA; Broad Institute of MIT and Harvard, Cambridge, MA, USA; Harvard Data Science Initiative, Harvard University, Cambridge, MA, USA; Kempner Institute for the Study of Natural and Artificial Intelligence, Harvard University, Cambridge, MA, USA

## Abstract

Electronic health records (EHRs) contain multimodal data that can inform diagnostic and prognostic clinical decisions but are often unsuited for advanced machine learning (ML)–based patient-specific analyses. ML models and clinical heuristics learn generalizable relationships from predefined factors, yet many patients may not benefit if those factors are missing in the EHR or differ—however subtly—from typical training populations. Clinical heuristics are limited to low complexity, often linear, relationships and patterns between clinical variables. ML approaches in EHRs significantly expand pattern sophistication but require large, labeled datasets, which are often unattainable especially in low prevalence diseases and are limited by sources of random and non-random variation in EHRs. Deep learning (DL), in contrast with ML and clinical heuristics, learns features without predefinition but requires even greater label access for predictions. While DL can construct unsupervised EHR representations, the patterns and characteristics of less prevalent examples are poorly resolved, and downstream clinical applications still require labels. We present Inf-EHR, a framework to automatically compute clinical likelihoods from whole EHRs of patients from diverse clinical settings without need of large volumes of labeled training data. We apply deep geometric learning to EHRs through a novel procedure that converts whole EHRs to temporal graphs. These graphs naturally capture phenotypic temporal dynamics leading to unbiased representations. Using only a few labeled examples, InfEHR computes and automatically revises likelihoods leading to highly performant inferences especially in low prevalence diseases which are often the most clinically ambiguous. To demonstrate utility, we use EHRs from the Mount Sinai Health System and The University of California, Irvine Medical Center and test its performance compared to physician-provided clinical heuristics across two diseases with no clinical or epidemiological overlap: a rare disease (neonatal culture-negative sepsis) with prevalence of 2% in neonates, and a more common disease (adult post-operative acute kidney injury) with prevalence of 22%. We show that Inf-EHR is superior to existing clinical heuristics both for culture-negative sepsis (sensitivity: 0.65 vs .041, specificity: 0.99 vs.0.98) and post-operative acute kidney injury (sensitivity: 0.72 vs 0.20, specificity: 0.91 vs 0.97). We present the first application of geometric deep learning in EHRs that can be used in real world clinical settings at scale, for improving phenotype identification and resolving clinical uncertainty.

## INTRODUCTION

Practicing evidence-based medicine is hindered by clinical uncertainty [1]. For any condition transitions from high-evidence regions to low-information, high-uncertainty areas are abrupt [2]. Clinicians must synthesize diverse sources of information to limit this uncertainty and make effective decisions [3]. Determining which information to use and how to combine it to estimate probabilities for decisions is complex [4] and relies heavily on individual judgment and experience, which runs counter to the principles of evidence-based practice [5].

This challenge is complicated by additional issues. First, clinical tests and heuristics generally have low positive predictive values and higher negative predictive values, necessitating multiple tests and extended clinical observation to reduce uncertainty [6]. In acute settings, this time delay is often weighed against the potential harms of empirical treatment and the risks of not providing it [7]. In more defined settings, such as pre-operative risk assessment, the window to gather and synthesize information is limited, establishing a baseline uncertainty that current heuristics and evidence struggle to resolve [8,9].

Electronic Health Records (EHRs) have evolved over the past decade, becoming increasingly detailed [10] due to technological advancements and changing regulatory requirements [11]. However, information contained within EHRs cannot readily be applied to individual clinical decisions [12]. While information rich, distilling EHR information into useful evidence requires identifying exactly which variables to look at and estimating the conditional relationships between them over evidence that varies across individuals and is subject to measurement and other errors [13]. Adding to this complexity is choosing the right temporal window in which to observe these variables for making a phenotypic inference [14].

Realizing the potential of the EHR to resolve clinical uncertainty requires novel computational approaches [15]. Graph structures, which consist of nodes connected by edges, are well suited to representing phenotypic relational structures [16]. Temporal structures are captured in the form of edges between nodes (representing clinical events) and the flexibility of this structure seamlessly captures individual variation (e.g. a node representing a certain medication may only selectively appear according to who received it) [17]. The graph structure compactly renders the clinical trajectory into a form suitable for AI applications [18]. Deep geometric learning is a highly active and emerging branch of AI research which, though nascent, has already produced impactful results in the biomedical domain [19, 20,86, 87, 88, 89]. Broadly, deep geometric learning extends advancements in neural networks to the case of data that consists in relational structures between entities (geometric data). In our specific application we use deep geometric learning to render EHR information into a clinical likelihood.

We minimize human involvement throughout the steps involved. This allows InfEHR to learn revealing temporal dynamics without human pre-specification or bias. Our method first automatically extracts temporal graphs from individual EHRs (EHR graphs). Each graph captures the complete trajectory of a patient’s clinical events and the relational structures between them. We summarize the entire clinical history contained in the EHR graph into a compact vector representation using self-supervised learning. This creates a holistic view of the patient record that expresses the semantic similarity between clinical trajectories as a function of spatial distance between vector representations.

This can be used to transmit knowledge to unlabeled cases from scant expert-provided labels as presumptive labels. We use these initial labels to refer to the EHR graphs and find individual graph components (e.g., a medication connected to a lab result) that are preferentially associated with an outcome, each providing a weak and uncertain likelihood estimate. We find hundreds of such informative components automatically and aggregate their weak predictions into refined machine derived likelihoods. These likelihoods provide statistical information that InfEHR combines with deep geometric learning to get a final result. The initial EHR graph is transformed by coalescing and connecting individual clinical events into learned higher order concepts (a learned graph), successive processing layers render the learned temporal graph into a likelihood according to a specialized training objective.

Our framework performs in criterion important for clinical applications. InfEHR computes likelihoods specific to individual cases for any patient. It does not rely on the presence of specific information such as a particular lab result or vital measurement. It also removes the requirement for a clinician to know when a model can be validly applied e.g., a model assumes only certain pharmacological exposures and becomes unreliable otherwise or is only valid in certain clinical settings. Implicit assumptions in the distribution of the training data may also invalidate a model. Even under valid conditions discriminative models can still report high confidence but unreliable likelihoods for uncertain cases. In contrast, the likelihoods returned by InfEHR naturally reflect its certainty in the individual case, indicating to the clinician exactly when there is not enough information in the EHR for it to make an inference. We observe accuracy to scale with confidence for both positive and negative predictions in all experiment settings, however, there are still cases over which InfEHR makes low entropy predictions (relative equiprobability between classes). As we will later discuss, such likelihoods may also express important phenotypic information concerning the instant case and are therefore still valuable in clinical decision making.

## RESULTS

### Uncertainty in Diagnosing Neonatal Culture-Negative Sepsis

The onset of inflammatory symptoms consistent with sepsis prompt empirical treatment with antibiotics [28]. Following a positive blood culture result, the criteria for antibiotic cessation are established by individual clinical response and other prior knowledge concerning the infecting pathogen. However, timing antibiotic cessation in cases without positive results and ongoing symptoms poses a challenge. Blood cultures, the most specific tests, involve processing times with incubation periods of up to 72 hours, and occasionally repeated samples to obtain reliable results [44]. Factors including maternal exposure to antibiotics and low sample volumes also contribute to clinical skepticism of blood culture results, adding to a period of diagnostic ambiguity while awaiting confirmatory results. In other cases, such as CN-S, confirmatory results will never come.

In these situations, a clinician must decide how to interpret the negative result; specifically, whether the patient has Culture-Negative Sepsis, a condition in which there is underlying septicemia requiring continued antibiotics or is experiencing inflammatory symptoms without an infectious cause [33]. Failure to treat sepsis is an unacceptable risk, however overuse of antibiotics is also associated with significant short and long-term adverse outcomes [34, 35]. Paradoxically, use of empiric antibiotics is also reported to increase risk of sepsis, NEC, or death (OR, 1.24; 95% CI, 1.17-1.31) [86]. The lack of consensus definition of CN-S or established clinical guidelines for its treatment complicates this situation and leads to highly idiosyncratic treatment patterns that are poorly supported by evidence [36, 37].

There is no known singular biomarker or group of biomarkers that specifically identify CN-S, and there is also no established criterion for pathogen identification based on clinical signs or measurements outside of a blood culture [40]. In neonates, rapid growth and development of organ systems also add variation to lab and vitals measurements which makes it difficult to independently discern a disease process [38] and many non-infectious conditions also lead to elevated inflammatory markers in neonates despite no underlying infection [41]. Serial observations and measurements over time are needed to form more cohesive clinical pictures [28] meanwhile the harms from inappropriate antibiotic exposure accrue. Exactly what evidence supports a CN-S diagnosis, and therefore continued antibiotics in the setting of a negative blood culture, is subject to ongoing debate [33]. In our sample of 8015 antibiotic courses (from 6596 individuals) an average of 72 (std. 32) unique labs excluding blood cultures are measured with a mean 225 (std 369) total lab measurements per course (mean course duration 4.35 days), however there is no consensus guidelines on how to use this information to resolve diagnostic ambiguity.

Temporal considerations add to the lack of consensus and diagnostic uncertainty. While several publications suggest the use of C-Reactive Protein levels to identify CN-S cases, they recommend measuring it at differing times from the start of antibiotics (from 3 days to 7 days at the earliest) [42, 43] as well as threshold level. As symptoms persist diagnostic uncertainty increases as the interpretation of a negative culture result becomes increasingly ambiguous. To model this and show the feasibility of InfEHR to reduce clinical uncertainty in realistic settings we extract EHR windows including 24 hours of information prior to antibiotic administration and up to and excluding the penultimate dose. The resulting windows reflect the patient’s clinical condition prior to the decision point to cease antibiotics.

We compute the conditional probability of a diagnosis (over all labeled conditions, see Supplementary Materials) given only elapsed time to find that by day 7 the probability of a CN-S diagnosis rises to 0.19 from less than 0.03 at day 3, approaching parity with an ROS diagnosis (Pr=0.27) reflecting high clinical uncertainty in the setting of ongoing symptoms with no positive culture result. The probability of an ROS diagnosis declines rapidly with time, while the probability of other diagnosis increases. We identify elapsed day 11 as the maximal window where all labeled conditions, including ROS, have non-zero probabilities. We therefore truncate all EHR windows to a maximum of 11 days to show that InfEHR can compute accurate likelihoods when multiple possible sequences are present. We find that the accuracy of InfEHR, as we will detail later, is mostly independent of the length of the underlying EHR window (spearman coefficient = -0.17, r squared = 0.031 p<.05).

We also found no correlation between truncation and accuracy, but truncated CN-S sequences were more likely to result in high entropy likelihoods (> 31% of truncated CN-S sequences had likelihoods between 0.4 and 0.6 compared to 9% full length) reflecting that while InfEHR effectively reduces uncertainty on incomplete sequences more information from longer observations improves prediction confidence.

An important point must be made about outcome label fidelity. There were 3657 EHRs corresponding to unique antibiotic courses manually labeled by a physician subject matter expert. Following the lack of consensus definition for CN-S the SME labeled a case as positive if the treating physician maintained the antibiotic course because of an intention to treat for CN-S. Unfortunately, due the lack of objective requirements for identifying CN-S there is possibility that some patients, despite being treated for CN-S, did not actually have CN-S. While it is not possible to directly identify these cases, if they exist, we hypothesize that they may comprise a subset of the cases with high entropy likelihoods.

### Uncertainty in Assessing Postoperative Acute Kidney Injury Risk Preoperatively

While static variables such as age, sex, and surgery type have been independently linked to PO-AKI [27], these associations and their interactions seem to be mediated through individual factors which vary by clinical setting [45]. This limits the use of generalized scoring indices or models in determining risk [46]. Here we express the concept of risk as the likelihood of developing PO-AKI and accordingly compute the prognostic likelihood of developing PO-AKI. We also depart from other methods by considering only time varying attributes of EHRs [54]. Our prognostic likelihoods are therefore dynamic in nature and can be modified by evolving EHR information. The likelihood corresponds to a patient at a given time subject to change with modifications to risk that occur throughout the clinical trajectory e.g., increasing drug administration (in contrast to static risk indices).

The link between surgery type and uncertainty of PO-AKI transmits along two lines: variation in the clinical setting comes with differences in the detail of monitoring and inherent perceived risks of PO-AKI and the medical background, including age, of a surgical patient [27]. Cardiac patients are routinely assessed for such risk following high incidence of PO-AKI for these patients (40% of all cases) [47]. Age has been identified as an independent risk factor in this setting; however, some cardiac surgeries seemingly carry greater risk than others and it is not entirely clear how risks stemming from age relate to this [48]. Nonetheless, numerous models exist for computing risk scores in the cardiac setting (with variable success rates) [49]. Non-cardiac surgery by comparison includes a more medically diverse group of individuals going through various types of surgeries with attendant differences in surgery lengths and other surgery-specific risk factors [45]. The SPARK index, acknowledging this intrinsic uncertainty, attempts to create a generalized clinical risk index for this population [54]. The index considers factors including pharmacological factors such as RAAS blockade, and physiological factors such as diabetes mellitus status and anemia [50, 54]. The performance of SPARK has been shown to degrade in populations with differing characteristics compared to the training cohort despite good performance in training and validated discovery cohorts [46]. While one study documents the performance degradation, there may be other unidentified cases and it is not obvious to a clinician if an instant patient is well covered by SPARK or not without detailed analysis over the alignment of a patient to the training distributions, a clinically infeasible possibility [52, 53]. This underscores a general concern with clinical models, including indices, in that it is not always clear how well they fit an individual patient despite ostensibly satisfactory population level performance [52].

The general response to better deciding individual risk has been to produce increasingly specific models [55]. We highlight the limitations of this approach by showing the full range of all possible lab measurements for patients within the dataset. Some lab measurements cover no positive cases while others are measured in clinical settings where PO-AKI risk is already high (i.e., most measured cases were PO-AKI Positive) and so it is unclear what additional information the lab may provide [56]. In settings where the pre-measurement probabilities were less biased, the number of individuals with the measurement can be low [57,58], an example of informed missingness leading to bias by indication [90]. Of the labs without frequency preference for PO-AKI positive or negative patients, no lab has an overall measurement rate above 32% and some labs, such as Prothrombin mutation, with less than 1%. Our dataset also consists of a medically diverse population spanning diverse age ranges (min=18, max=92) undergoing surgeries over every physiological system (excluding reproductive systems) [27]. This comprises a group of individuals with collectively high clinical uncertainty that is challenging to resolve using a single model despite established risk assessments for individual subsets [52].

38.7% of our dataset had pre-surgical eGFRs below 60, of those only 37.1% developed PO-AKI showing that substantial uncertainty remains even after establishing baseline estimations of kidney function. InfEHR can include information even from labs with low representation, along with other factors facing similar measurement related constraints such as vital information or medications provided by finding commonalities across settings and holistically integrating individual information to resolve uncertainty. We assigned positive labels to all patients with AKIN scores > 0 at 72 hours post-surgery and negative labels otherwise to design a trainable task. However, we report the likelihood of developing PO-AKI using only pre-operative data. For a subset of positively labeled patients with low-likelihoods returned by InfEHR, it is possible but unknown if the factors involved in developing PO-AKI occurred primarily during surgery or immediately following the operation. In this case the model output would be correct where we currently deem these cases as failures in recall. Similarly, where InfEHR was uncertain, it is possible that there was some risk enhancement but unpredictable events in surgery that ultimately led to AKI. Unfortunately, we are unable to discern these patients from pre-operative data. These are limitations to predicting outcomes pre-operatively where in-surgery events may modify the outcome.

#### Performance of clinical heuristics under conditions of uncertainty

Diagnostic complexity and the constraints of clinical practice limit the range of information a practitioner can use in medical decision making [59]. Heuristics, or decisional shortcuts, are an adaptive strategy to efficiently manage uncertainty under time constraints [60]. This consists of integrating pattern recognition based on experience with pathophysiological reasoning to identify the most salient aspects of the instant case [61]. These cognitive operations resemble clinical tests in that they also have implicit pre and post-test probabilities (typically unknown to the practitioner) [4].

Here we obtain such heuristics from practicing physicians and apply them over the empirical distribution of cases to quantify the implicit uncertainty involved in using them. We corroborated the physician-provided heuristics with literature to ensure that they are representative of general practice and not limited to institutionally specific patterns of care. We apply the heuristics to the EHR windows as described above to compute likelihoods at clinically relevant points of care and examine their probabilistic outputs over the empirical distributions of cases in each dataset.

To quantify the performance of the heuristic we compute the rule-in and rule-out potentials [62]. Rule-in and rule-out potentials are prevalence-independent metrics based on the characteristics of probability distributions produced by a clinical test. They quantify a diagnostic test’s innate capacity to revise disease probability before testing occurs. Unlike likelihood ratios, which only characterize performance after a specific test result is known, these potentials predict how effectively a test or heuristic will revise disease probability for an average subject prior to performing the test. A rule-in potential of 2.0 indicates diseased subjects are, on average, twice as likely to be correctly identified after testing, while a rule-out potential of 2.0 means non-diseased subjects are twice as likely to be correctly excluded. Together the rule-in and rule-out potentials express the information gain provided by a test relative to the two poles of clinical decision making [64]. Rule-in and rule-out potentials capture a test’s inherent discriminative power rather than just its observed performance on a particular dataset (including prevalence differences). We additionally report sensitivity and specificity which are also prevalence independent [66] (see Table 1). While these metrics do not capture probability revision capacity or information gain, they do measure performance within already-known groups (diseased/non-diseased) providing an additional view on a test’s innate capacity to confirm disease presence. While we emphasize these dataset independent measures, we also plot the cumulative probability density functions by disease status (see Figure 2 and Figure 3) to illustrate the performance distribution across our specific datasets.

**Table 1:** InfEHR outperforms clinical heuristics across disease settings.

|  | Rule In | Rule Out | Sensitivity | Specificity | PPV | NPV |
| --- | --- | --- | --- | --- | --- | --- |
| **CN-S** |  |  |  |  |  |  |
| Heuristic | 1.013 | 1.0097 | 0.041 | 0.983 | 0.31 | 0.85 |
| GNN | 14.139 | 2.736 | 0.65 | 0.994 | 0.802 | 0.986 |
| **PO-AKI (MSHS)** |  |  |  |  |  |  |
| Heuristic | 1.309 | 1.154 | 0.197 | 0.978 | 0.695 | 0.824 |
| GNN | 3.269 | 2.455 | 0.723 | 0.911 | 0.678 | 0.927 |
| **PO-AKI (UCIMC)** |  |  |  |  |  |  |
| Heuristic | 1.326 | 1.121 | 0.13 | 0.993 | 0.717 | 0.904 |
| GNN | 6.497 | 2.551 | 0.65 | 0.978 | 0.787 | 0.95 |

**Figure 1:**
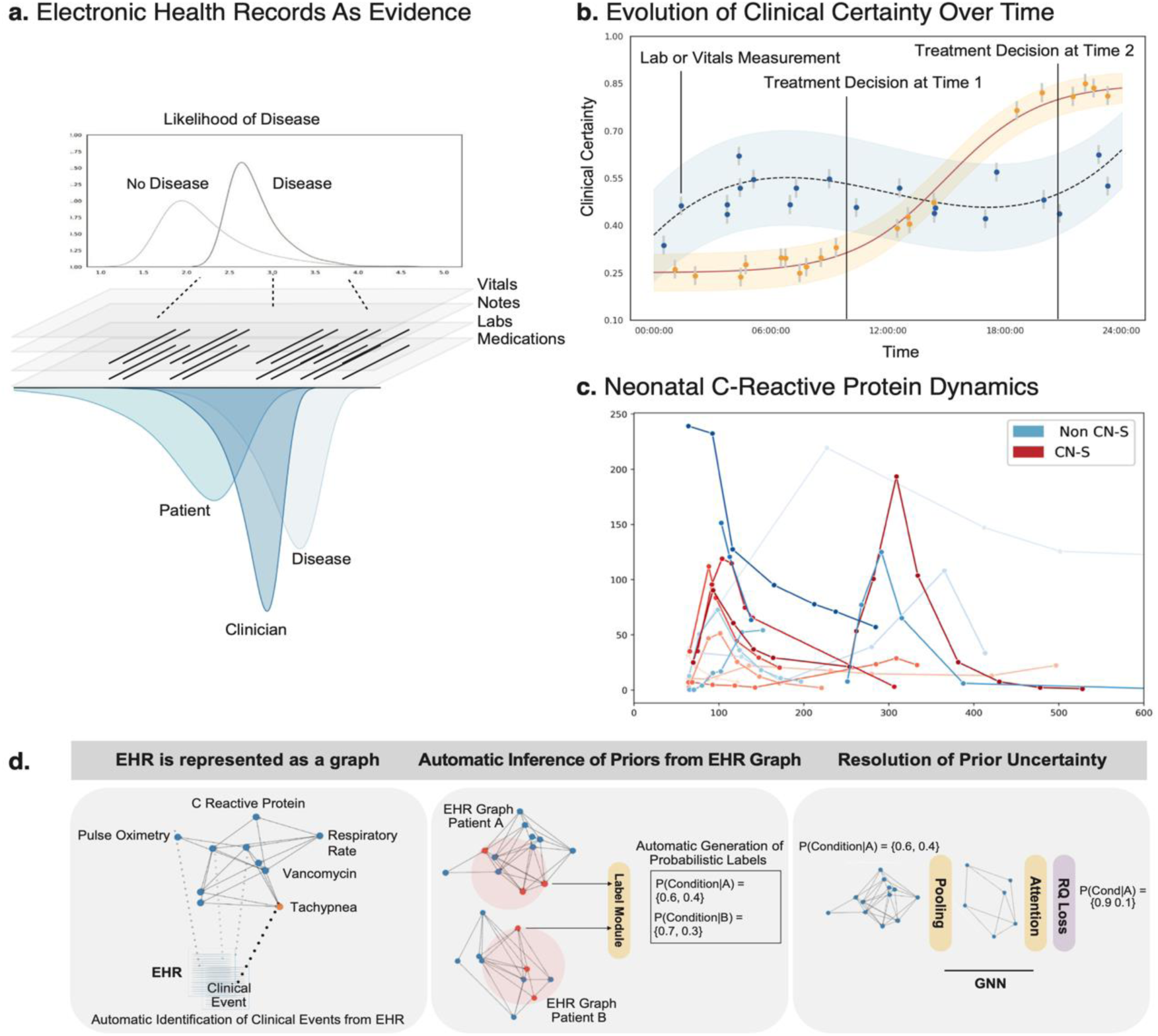
InfEHR is a framework for resolving clinical uncertainty using probabilistic and deep geometric learning on Electronic Health Records (EHRs). A. An underlying disease is only one of several generative processes evidenced in an EHR. The EHR results from the interaction between a disease state, its presentation in a particular patient, clinician response and non-clinical factors. Algorithms that do not consider these generative processes are challenged to make reliable inferences from EHRs. In contrast, InfEHR integrates these dynamics into its computed likelihoods achieving superior performance under clinically realistic conditions. B. Clinical certainty dynamically evolves non-linearly over time according to the patient. For patients well fit to existing clinical knowledge, low certainty states can be improved upon accumulation of the right information (in yellow), however for a more clinically ambiguous patient (in blue, wider uncertainty estimates), existing heuristics and clinical tests may not provide added clarity leading to highly uncertain decisions or delay pending more information. C. Despite the intuition that temporal dynamics provide distinguishing phenotypic information (e.g., CRP level dynamics in CN-S) this information is difficult to extract and apply in the clinic. d. InfEHR solves both problems by making holistic use of EHR information including temporal dynamics. We show performance in the setting of a rare disease with no clinical consensus definitions (Neonatal Culture Negative-Sepsis) and in predicting a more common condition (Post-Operative Acute Kidney Injury) for which the estimation of likelihoods is reportedly challenging.

**Figure 2:**
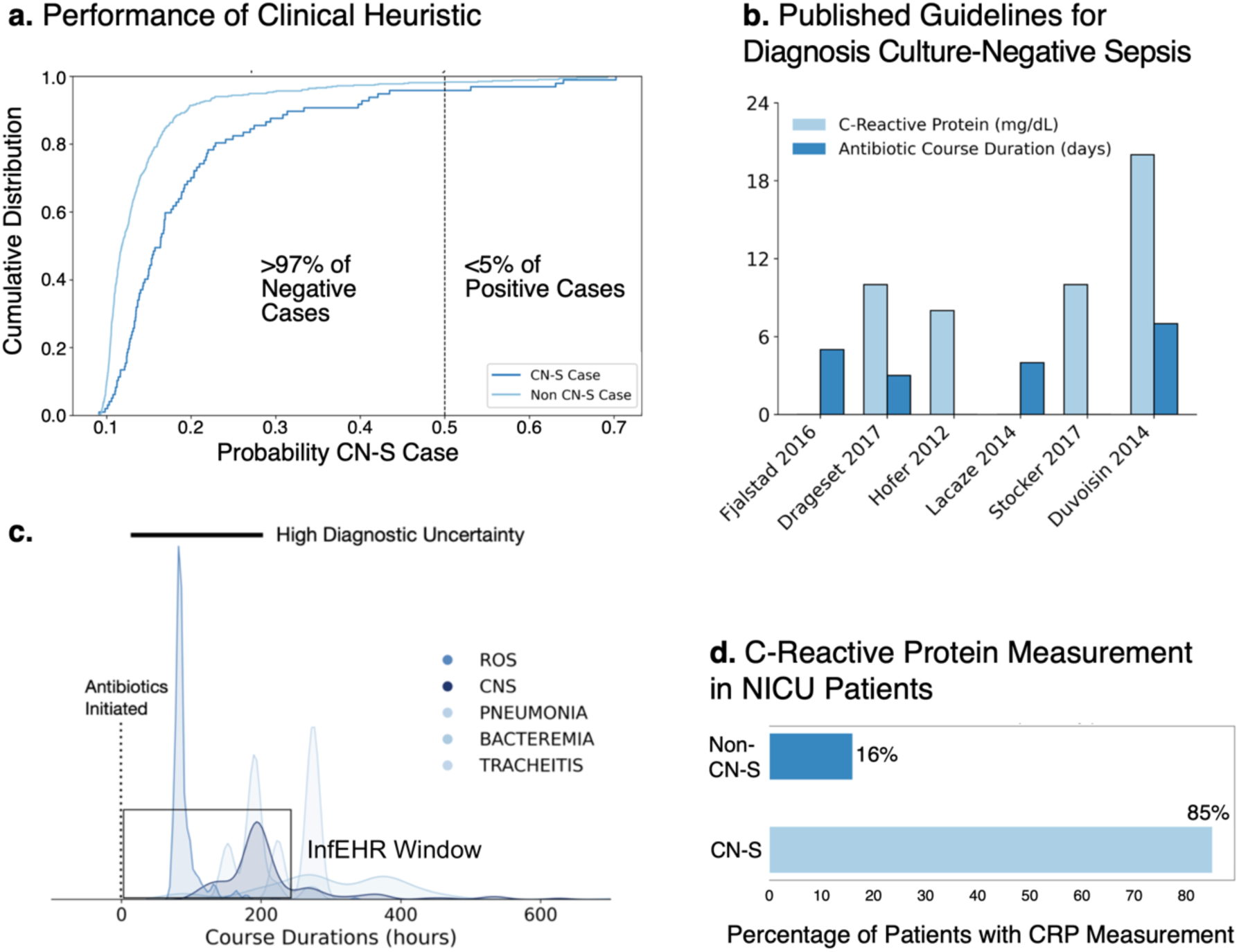
Clinical uncertainty in the setting of Neonatal Culture-Negative Sepsis: Antibiotics are administered empirically to patients presenting with systemic inflammatory symptoms, deciding when it is appropriate to stop is clinically uncertain. Inflammatory symptoms can persist despite negative culture results in certain patients. Rarely this correlates with septicemia despite a negative culture result. Meanwhile continuation of antibiotics accumulates significant risk of co-morbidities. **A.** We apply a clinical heuristic for identifying CN-S cases provided by a physician expert and corroborated by literature. The heuristic correctly identifies 97% of negative cases but fewer than 5% of positive cases. **B.** There is no consensus clinical definition for CN-S with guidelines variably suggesting time points with varying (including no) C-Reactive Protein level criteria. Clinical decision making for antibiotic continuation is therefore made with little information or guidance. **C.** Ongoing inflammatory symptoms without confirmatory culture results prompt continuation of antibiotics. This clinical state is temporally consistent with CN-S, other infectious conditions, and conditions that do not require antibiotics. InfEHR provides highly accurate likelihoods using the EHR information limited to this window. **D.** Despite some suggestions that CRP can help identify CN-S, it is not always measured in patients with CN-S. 15% of patients without CN-S had CRP measurements in our cohort however they significantly outnumber CN-S patients. These factors collide with ambiguity over measurement timing and thresholds to limit the value of CRP in reducing uncertainty. InfEHR does not solely rely on or require CRP measurements. When CRP is measured, InfEHR contextualizes the values to the whole EHR (including measurements over time) maximizing information in CRP for uncertainty reduction.

**Figure 3:**
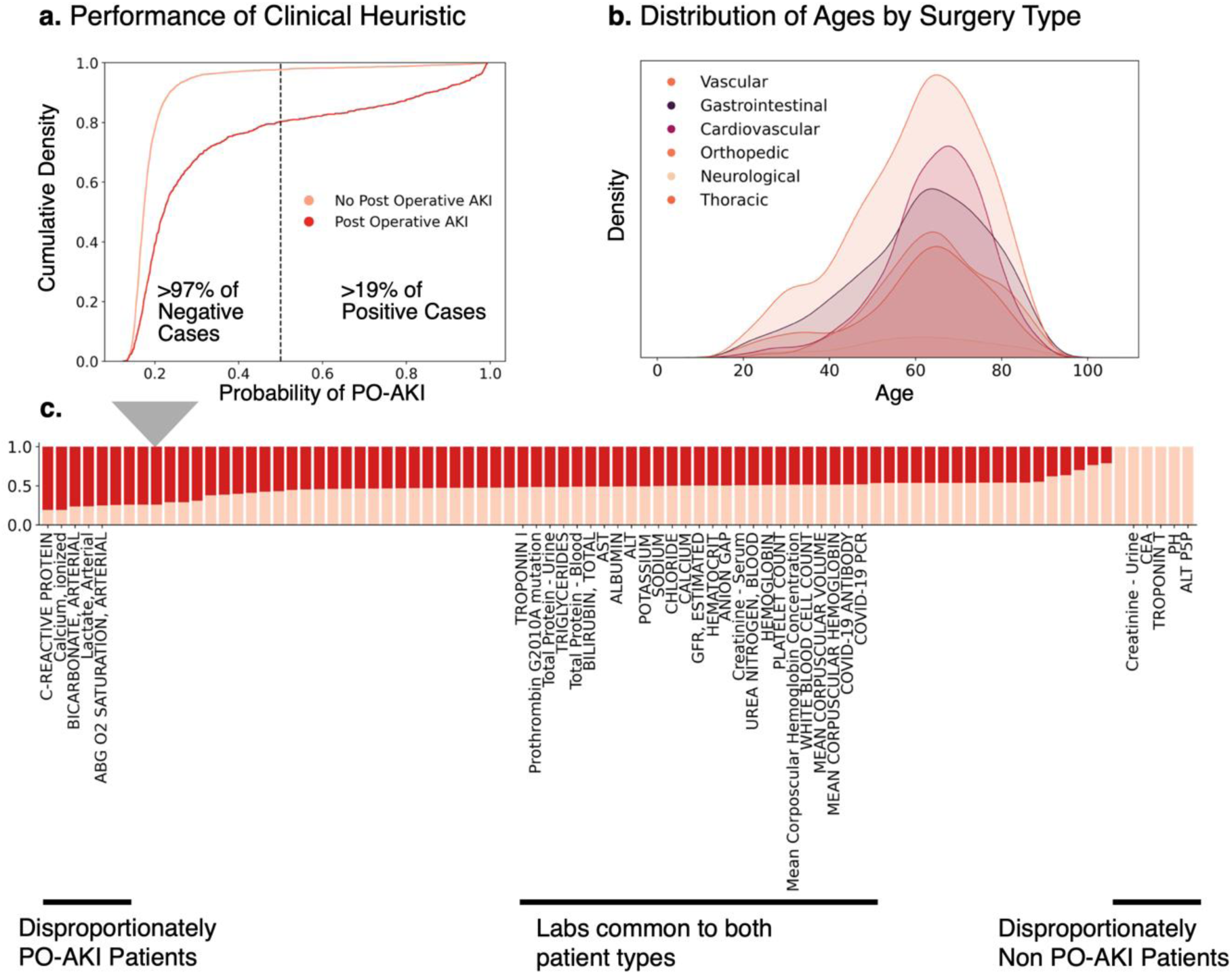
Clinical Uncertainty in the setting of predicting Post-Operative Acute Kidney Injury (PO-AKI) pre-operatively. Acute Kidney Injury leads to short- and long-term adverse outcomes that progressively worsen with interventional delay. Many models predict PO-AKI, but the highly conditional nature of risk factors has led to models intended for specific clinical settings and populations, narrowing their applicability. **A.** We apply a physician provided clinical heuristic using serum creatinine which is widely measured (including for all patients in this dataset) across patients and settings. The heuristic identifies > 97% of negative cases but < 19% of positives indicating that more information is needed to identify these positive cases. **B.** Patients in this dataset comprise a diverse pre-operative cohort spanning wide age ranges and undergoing surgeries of various types. PO-AKI risk is concurrently variable and dependent on individualized factors. **c.** Many labs, while informative, are not commonly measured across clinical settings or patients which limits their generalizability (e.g. urine protein). Models predicting across settings are typically limited to labs or other measurements common to each patient. InfEHR overcomes this limitation and computes individualized likelihoods, automatically using all relevant available information (including vitals, notes, and meds). Consequently, InfEHR identifies significantly more positive cases compared to the heuristic and can also generalize to diverse patients and settings.

##### CN-S Heuristic

Single point assessments of clinical signs have little value in diagnosing CN-S as multiple sources of variation in the perinatal environment underlie the observed signs [67]. No uniform guidelines exist for serial measurements or their interpretation [28]. Biomarkers, including Inflammatory biomarkers, generally have low sensitivity and specificity for CN-S, but some literature supports some positive predictive value for C-Reactive Protein (CRP) [42]. Accordingly, we use a clinician provided heuristic using the minimum, maximum, and average CRP concentration within the prediction window. CRP levels have been shown to vary according to gestational age, however including this information did not improve heuristic performance [43]. CRP is not uniformly measured in all CN-S cases (18% of CN-S cases had no CRP measure) [32]. The rule-in and rule-out potentials indicate that the CRP based heuristic did not reduce uncertainty (rule-in potential = 1.013, rule-out potential = 1.001). It is possible that specific measurement timings or other parameters about CRP measurement may enhance the clinical value of CRP in diagnosing CN-S, however there are no consensus guidelines to these parameters. This ambiguity contributes to the low diagnostic potential and an NPV of 0.85. The ECDFs indicate some separation between the likelihoods by class suggesting that CRP potentially holds more diagnostic information than our heuristic, and as we estimate a clinician on average, could extract.

##### PO-AKI Heuristic

Serum creatinine is commonly measured across various pre-operative settings (100% measurement rate for patients in the dataset) and is also used in validated predictive models or indices for PO-AKI in cardiac and non-cardiac surgery [45, 49]. We are unaware of any clinical signs validated for use in PO-AKI prediction and therefore did not consider any for heuristic use. Of the remaining 100 labs common to all patients in the dataset (less than 21% of all labs measured) none were recognized by a physician as having added prognostic value. We include length of stay measured in fractional days along with serum creatinine since this enhanced heuristic performance by serving as a non-specific index of case severity [68]. We use mean serum creatinine, last pre-operative serum creatinine and length of pre-operative stay to predict PO-AKI (AKIN=3).[27] The returned likelihood can be considered as a risk-index [69]. This heuristic has a rule-in potential of 1.309 and rule-out potential of 1.154. The heuristic provides some uncertainty reduction especially with some positive cases but there remains a subset of positive cases which the heuristic was unable to identify (NPV = 0.85, and as seen in the ECDFs figure 2).

The heuristic performances illustrate recurrent challenges in identifying positive cases. (PPV = 0.67, FNR=0.79) While it is apparent that additional information is required to resolve uncertain likelihoods exactly which additional information and how it can be integrated is unclear. It is also unclear to a clinician when a heuristic may not apply to a given patient even if overall performance is otherwise known. In our examples, false negatives can be explained by a general lack of discriminatory capacity of the heuristic (CN-S) but also by poor fidelity of an otherwise performant heuristic to the individual case (PO-AKI). As we explain in more detail next, InfEHR both automatically creates heuristics built on the particularities of individual cases as well as integrates the array of generated heuristics into a single likelihood that can later be further resolved to enhance rule-in and rule-out potentials.

### Automatic Generation of Heuristics from EHR Graphs

We use an automatic process to transform individual EHRs into temporal graphs where clinical events are identified and represented as nodes and connected to each other by edges according to temporal relationships in the record (detailed below). We use the graph structure to generate the automatic heuristics in a two-step process. First, a GNN, trained on an unsupervised objective, encodes the EHR graph into a compact numerical representation. The representation is designed to capture distinguishing features of an EHR graph (such as clinical event composition, or the existence of certain temporal dynamics) and assign them to a coordinate in a high dimensional semantic space (the representation). The resulting coordinate space aligns graphs (generated from patients) according to similarities in distinguishing features. By taking the sample of expert provided labels (110 total) and fitting a distance-based label propagation algorithm, we can presumptively label cases by a learned spatial distance metric [70]. This provides an initial set of labels for all the records according to a structural and spatial view of similarity between individuals as expressed through their encoded graph representations. We further evaluate the self-supervised representations quantitatively by analyzing the nearest neighbor of each representation in terms of ground truth label expressed by Mean Average Precision. The score, ranging between 0 and 1, reflects the percentage of times the nearest neighbor for a given representation shares the same ground truth label (MAP@1 CNS: 0.79, MAP@1 PO-AKI: 0.71).

We use the presumptive labels from the spatial representations to derive new labeling heuristics. We randomly select positive and negative cases from the 110 member sample. Next, we identify highly connected nodes within the EHR graphs corresponding to these selected cases and extract the 1-hop neighborhood of such nodes. Given that EHR graphs consist of observed relationships between clinical entities (expressed as edges between nodes), the resulting extracted subgraph then contains potentially identifying clinical relationships. If the relational structure preferentially distributes in EHR graphs from a given label, we use the relationship to heuristically assign labels. Therefore, we use elements of the graph structure itself as weak labeling heuristics.

To identify these structures, we use the labels obtained by spatial propagation over the high dimensional representations of the EHR graphs (as described above) to assign labels to individual nodes. We assign the node label according to its class association. We can identify relational structures from the sub-graph using these labels according to a simple procedure: when two or more connected nodes share the same label, we treat the substructure as a labeling heuristic which assigns the common node label to any graph where it is present. To improve the accuracy of these weak heuristics, we check if any of the connected nodes are also connected to nodes of opposite label. In this case, we remove any individuals identified by both the same label and opposite label substructures from labeling by the heuristic, limiting application to cases without known contradictory information.

We generate 2400 negative and 396 positive labeling heuristics for CN-S and 4307 negative and 982 positives for PO-AKI. We observed that many such automatically generated heuristics are corroborated by findings in literature (see Supplementary Materials for specific examples). Despite filtering for contradictions within heuristics (e.g. an individual is predicted to be both positive and negative), there remain substantial contradictions between heuristics. However, the heuristics can still be effectively combined following Ratner et al. [71] to produce performant likelihoods. These likelihoods represent coalesced information from whole EHR-to-EHR comparisons (i.e., labels derived from the self-supervised embeddings of EHR graphs) and specific conditional relational structures (the weak labeling heuristics). To improve the information in the likelihood distribution we obtain the final likelihoods by randomly subsampling and combining 100 positive and 100 negative labeling heuristics. We repeat this process until there is only minimal change in the average entropy of the resulting averaged likelihood distributions.

The resulting distributions are more performant in identifying positive cases compared to clinician provided heuristics in both disease settings (PO-AKI: FNR 0.48 vs. 0.79 clinician heuristic, CN-S: FNR 0.78 vs 0.95 clinician heuristic). The machine generated heuristics also retain uncertainty in their distributions (Empirical CDF Pr(0.2 - 0.8) CN-S: 0.32, PO-AKI: 0.43). This allows the GNN component of InfEHR to learn uncertainty reducing temporal dynamics in part by identifying cases with ambiguous label dispositions (high entropy label probabilities) and using features derived from low entropy cases to resolve them. This process would be significantly impaired by overly confident input distributions with low positive case recall (particularly in the setting of rare or infrequent disease). The distribution is used as prior information in the loss function of the GNN, which ultimately reduces individual entropy, as we explain below.

### Deep Geometric Learning Resolves Prior Uncertainty

The machine generated likelihoods derive from clinical relationships encoded into the structure of EHR Graphs, expressed here as a connection between two or more nodes each representing a clinical event. We expand the representational capacity of an EHR graph by including (1) semantic encodings for each clinical event as attributes (assigned to each node in the EHR graph) and (2) time stamp encodings which can be combined with the semantic encoding in (1) to create attributes reflecting individual temporal contexts (Figure 4). These attributes build on the existing temporal structure, encoded by the collected nodes and edges of an EHR graph, into a format suitable for learning phenotypic temporal dynamics at scale. The semantic encodings in (1) automatically reproduce clinical knowledge in terms of spatial distance, e.g. the nearest neighbors of an encoding for a certain respiratory rate include representations for pulse oximetry. We also observe less established but nonetheless corroborated clinical relationships such as the appearance of encodings for eosinophil measurements within the near neighborhood of ejection fractions. The detailed and individualized consideration of the semantic and temporal information added to the EHR graphs through these encodings is executed by training a GNN to return likelihoods from individual EHR graphs using the previous machine-generated likelihoods as priors to be resolved.

**Figure 4:**
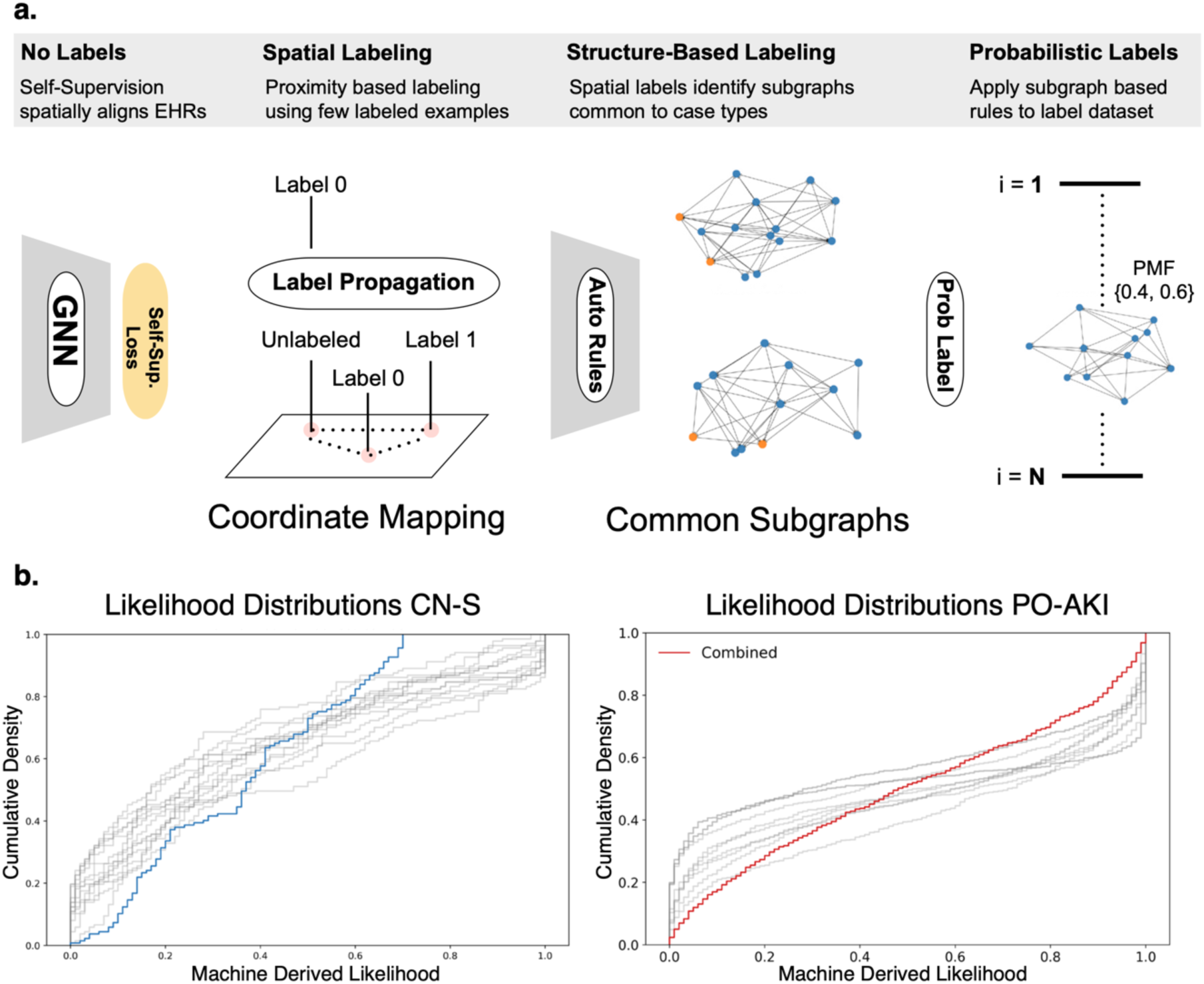
Initial uncertainty resolution through automatic probabilistic labeling. InfEHR uses self-supervised learning in combination with graph derived weak heuristic predictors to automatically derive likelihood for unlabeled cases from sparse labeled examples. **A**. EHRs are encoded to compact representations in a coordinate space that balances individual representation while preserving semantic relationships between similar patients. Similarity between individuals can be measured by spatial proximity which is used to apply presumptive labels to unlabeled cases. Subgraphs are extracted from labeled cases and used to generate programmatic labeling rules or heuristics that combine topological information from the EHR graphs with spatial information from their compact encodings into probabilistic labels for the entire dataset. We repeat this process and average the results to obtain initial individual likelihoods (in blue and red). **B**. The resulting distributions indicate significantly improved ability to identify positive cases (> 35% CN-s, >50% PO-AKI) while maintaining discrimination of negative cases (>92% both settings). InfEHR resolves remaining uncertainty using the automatically generated probabilistic labels as priors and performing probabilistic revision through deep geometric learning of phenotypic temporal and structural features in the EHR graphs.

We train the GNN under a specialized loss function[72] designed to compute posterior likelihoods following a generative modeling framework. In consideration of the input structure over which the training is performed, this amounts to identifying temporal dynamics between clinical entities that are likely to be generated from an underlying disease latent. Since the clinical entities and their temporal orderings are extracted from EHRs, the prediction task can be seen as predicting the likelihood of an EHR under an assumed disease process (or physiological state). The GNN is trained end-to-end to learn the most phenotypically relevant subgraph and node representations primarily based on clinical entity types and then to learn the identifying temporal dynamics within this subgraph to determine a likelihood. While the parameters required for the graph distillation and likelihood computation are learned batchwise, the parameters are applied to individual EHR graphs and use the specifics of individual cases, in which we make no assumptions or requirements over shared clinical trajectories or backgrounds between patients, to compute highly accurate likelihoods.

The resulting likelihoods markedly outperform the clinician provided heuristics with superior rule-in and rule-out potentials in both settings. We highlight here that this required and potentially even benefited from no human input besides a small number of initially provided examples. The resulting rule-in potential for CN-S of 14.39 (GNN) compared to 1.013 (clinician provided) shows the GNN is about 14 times more likely to identify the average positive case compared to the clinician provided heuristic. This finding is also reflected in the PPV and NPV which are about 2.5 and 1.16 times greater than the clinical heuristic. In part this represents the extent of resolvable uncertainty within the clinical setting. The GNN produces comparable results in the setting of PO-AKI risk assessment with 2.5 times greater rule in-potentials and 2.13 greater rule out potentials. The GNN also reduces the FNR as reflected by 1.12-fold increase in NPV. This does come at a small expense of slightly higher false positivity, the GNN PPV performs about 2% lower compared the clinical heuristic while making higher overall true positive predictions (identifying 71% of all positive cases compared to only 19% by the clinical heuristic).

#### Validation of InfEHR on an External Dataset

We evaluate the performance of InfEHR on predicting PO-AKI using EHRs contained in the Medical Informatics Operating Room Vitals and Events Repository (MOVER, public credentialed access) [92]. This dataset includes a medically diverse population of patients undergoing surgical procedures at the University of California, Irving Campus Medical Center. We obtain n=2427 patients for whom we could both assign PO-AKI status (AKIN scoring system) and apply the clinical heuristic; of these patients we identify n=261 positive cases (∼10% prevalence vs. ∼23% Mount Sinai Health System (MSHS)).

We evaluate the InfEHR framework as follows. (1) We construct EHR graphs as previously described for all MOVER patients. We transfer knowledge from MSHS by aligning MOVER EHRs with MSHS and extracting clinical events as learned from MSHS data forming the nodes and setting as their attribute’s the embeddings obtained from the previously learned manifold. (2) The MOVER dataset does not contain clinical notes. In order to generate prior probabilities, we processed all MSHS graphs and ablated any nodes originating from note sources and then re-trained the GNN and obtained prior probabilities for each EHR graph in MOVER (the input distribution) resulting in a rule-in and rule-out potential of 4.32 and 1.96, respectively. InfEHR improves this input distribution to achieve rule-in and rule-out potentials of 6.34 and 2.52 respectively while reducing the FNR from 0.46 to 0.34 which shows that InfEHR is effective in automatically resolving uncertainties in prior distributions without human intervention. As we will discuss further this sets InfEHR apart from discriminative models (including traditional ML) and provides a significant advantage in clinical applications.

## DISCUSSION

Low disease incidence intrinsically limits the diagnostic power of common individual tests [73]. Similarly, underlying risk accumulation may proceed through highly conditional structures which can make individual risk difficult to assess since individual variation within those structures may lead to correspondingly divergent risks [74]. These factors, along with limitations in time and existing knowledge, promote uncertainty in clinical decisions [2]. Discriminative models, even under optimal configurations of model architecture and type, are poorly situated to resolve these uncertainties. The discriminative modeling approach consists in trying to predict a label given a data instance. Models trained under this framework learn to predict a label or class membership through incurring a penalty for incorrect predictions. As we will explain this approach is conceptually mismatched to the realities of EHR data and clinical medicine, especially in the setting of low prevalence diseases which are typically also the most clinically uncertain and where input from models is wanted most.

Discriminative models in medicine can fail when handling rare conditions due to dual challenges: sparsity in both the number of positive cases and their distribution in feature space [93]. With few positive examples, the model struggles to learn the true variety of ways a condition can manifest - it may only see a narrow subset of possible presentations and optimizes for learning a decision boundary primarily on the abundant negative cases. This limited exposure makes it difficult for the model to develop robust feature representations that capture the full spectrum of disease manifestations and the decision boundary itself becomes problematic because it is primarily shaped by the dense regions of negative cases, with only sparse positive cases to counterbalance this influence. This can result in the model making highly confident but incorrect predictions.

Cases that fall far from the learned feature space of the training data but definitively on one side of a decision boundary represent a fundamental flaw in discriminative models.[95]. The model will assign high confidence simply based on the case’s position relative to the boundary, even though we have little evidence to support such certainty in sparse regions of the feature space (here the model has seen few or no other cases with similar learned features). Sparsity in the feature space may be due to limited training examples (common in rare conditions such as CN-S) or in combination with poor inductive capacity of the model to learn useful features (e.g., highly conditional networks underlying PO-AKI risk). This creates a dangerous situation: a clinician reviewing the model’s output would have no way to know that the high confidence prediction comes from a region where the model has minimal experience.

Post-hoc calibration cannot solve this problem because it only adjusts probability outputs without addressing the underlying issue - the model’s restricted understanding of how diseases manifest due to the nature of discriminative training [94,96]. Consider a rare disease that can present in several distinct ways. If the training data only captured one type of presentation, the discriminative model builds its decision boundary around that presentation. When encountering a different but equally valid presentation of the same disease, the model might confidently misidentify it simply because the case falls on the "wrong" side of the boundary based on the learned features. Calibration techniques cannot fix this fundamental gap in the model’s knowledge because they can only work with the feature space (the way a case is ultimately represented within the model) that the model has already learned. The calibration process can’t teach the model about alternative disease presentations it has never seen or lead to output probabilities that reflect when a model is operating with epistemic uncertainty which severely limits the applicability of likelihoods from discriminative models in clinical settings even when calibrated.

In contrast, the generative approach explicitly models how different disease processes could generate various presentations in the EHR and simultaneously weigh the consistency of a given case with the presumption of a given disease. Generative models incorporate disease prevalence as a core component of this reasoning - rare conditions require stronger evidence to overcome their low prior probability, just as clinicians maintain a higher threshold for diagnosing uncommon conditions. The likelihoods returned by generative models critically preserve uncertainty in cases where the learned EHR features are not more consistent with any disease presumptions. Rather than making decisions solely on feature patterns, the model considers both how well the patterns match each disease process and how likely each disease is to occur. Deep geometric learning provides an opportunity for learning highly informative and patient specific EHR representations leading to well resolved feature spaces. When combined with a generative approach we are able to achieve a more nuanced and clinically valuable form of uncertainty quantification - a case might have features somewhat consistent with a rare disease, but the model appropriately and automatically tempers its confidence based on the condition’s rarity or the model may be inexperienced with an instant case leading to similarly moderated likelihoods.

We introduce InfEHR, a generative modeling framework, which we have shown to dramatically reduce clinical uncertainty of individual cases using EHR data available at the time of clinical decision making. Our framework automatically learns phenotypic temporal dynamics from EHRs through a graph neural net-based approach. We learn high-dimensional representations of such graphs to compute the likelihood of an underlying disease latent given the representation (and the EHR it was derived from). Our resulting likelihoods have properties important to any clinical test: the likelihood clearly identifies when the model is uncertain, confidence scales with accuracy, and the likelihoods substantially revise pre-test probabilities [78, 80]. We express these characteristics quantitatively through high rule-in and high rule-out potentials benchmarked against real world clinical heuristics. To our knowledge, we depart from existing deep geometric learning approaches to EHRs[21] by considering temporal graphs to (i) automatically derive clinically significant nodes and their embeddings from discrete and continuous variables within raw EHRs, (ii) use a graph structure in a rules generation engine to produce informative priors (allowing us to use minimal labeled examples), (iii) learn whole temporal graphs and representations from EHRs using semi-supervised and unsupervised deep geometric learning, and (iv) compute likelihoods from these representations for resolving clinical uncertainty in realistic conditions (e.g., we consider EHRs as they exist at the time of decision and do not rely on diagnosis codes or other structures that may not be available at any decision time).

We also demonstrate that our framework can be used to automatically revise prior probabilities generated from previously trained models applied to new data with differing underlying prevalence of disease. This sets InfEHR apart from discriminative approaches for a few reasons: (i) underlying differences in prevalence typically manifest as degraded model performance in discriminative models. This can be palliated by sufficient training examples covering all potential disease manifestations, but low disease frequency intrinsically limits the availability of these training examples. In contrast, InfEHR does not require large volumes of training data to prevent performance degradation since the method of its training explicitly models prevalence in its likelihoods. (ii) Fine tuning previously trained models to better reflect local prevalence conditions requires re-labeling an entire dataset which is often not possible. InfEHR instead revises prior probabilities automatically according to local prevalence conditions without any human labeling. (iii) InfEHR learns local feature distributions that can be combined with automatic inference of prevalence that lead to high rule-in and rule-out potentials which supports the generalizability of the framework to new settings with differing clinical practices and patient populations.

InfEHR has a wide range of clinical applicability. We compute likelihoods in disease settings with low (PO-AKI) and very low (CN-S) incidence of positive cases. While the true global incidence of PO-AKI is not presently known, a recent publication estimates that 18.4% of surgical patients will develop it (95%CI 17.7 - 19.2%) [27] while we observe 21% in our training dataset (n=4276, PO-AKI = 879) and 10% in our validation dataset (n=2426, PO-AKI=261). Estimates of global CN-S cases are more elusive however in our dataset CN-S cases make up less than 4% of the total cases (n=3678, CN-S=137) which is consistent with ratios available in literature [28, 29].

While we emphasize performance in positive case identification in low prevalence settings InfEHR is also performant in finding negative cases as it returns likelihoods with high rule-in and high rule-out potentials as we explain below [31].We show the performance of InfEHR in EHRs obtained from the Mount Sinai Health System (MSHS) on CN-S and PO-AKI, and validate the performance in PO-AKI prediction on EHRs from the University of California, Irvine Medical Center (MOVER). The EHRs in all datasets are from patients with diverse medical and demographic profiles [25]. As we explain in detail below, we also demonstrate the performance of InfEHR under clinically realistic conditions in terms of decision timing and EHR availability. This approach, requiring minimal human intervention, marks a significant advancement in leveraging EHR data to support evidence-based medicine and reducing clinical uncertainty [20, 21,26].

Identifying low-incidence cases is particularly challenging to clinicians and machines alike [30]. The absence of uniform diagnosis and management, such as in diagnosing neonatal CN-S (where there is no specific confirmatory result or set of results), compounds these challenges. We benchmark InfEHR against a clinical heuristic for CN-S identification that emphasizes CRP given that it is the only biomarker with specific guidelines for use in diagnosing CN-S [42]. Absolute CRP levels in neonates are limited in distinguishing physiologic from inflammatory response, as it is subject to natural developmental increases outside of inflammatory response [43]. Further, various non-infectious conditions in the perinatal period can also induce an inflammatory response such as complicated labor and delivery, intraventricular hemorrhage, or tissue injury [41]. These issues likely reduce the sensitivity of CRP in CN-S detection as the heuristic, consistent with literature, shows low sensitivity (0.041) and correspondingly neutral rule-in and rule-out potentials (1.03 and 1.097) [63]. In contrast, InfEHR dramatically improved upon these results with sensitivity of 0.650 and high rule-in and rule out potentials (14.139, 2.736). We observe recurrent and distinguishing CRP level temporal dynamics in some CN-S positive cases (Figure 1). This observation suggests value in considering temporal dynamics for separating physiologic from pathophysiologic responses [76]. However, identifying such dynamics, which are subject to individual variation within a conserved pattern, at scale exceeds the capacity of the heuristic and is challenging to traditional temporal models [91]. InfEHR automatically captures informative temporal dynamics without algorithmic or human pre-specification and can also consider interrelating variations between CRP and other biomarkers such as in CBC results which, by themselves, while reportedly low in sensitivity, may provide more information when integrated with other results.

The proliferation of models and clinical indices for the assessment of PO-AKI underscores the highly contingent nature of PO-AKI risk [55]. Patients with normal serum creatinine do not present with clear post operative risk and are highly represented in the false negative predictions (FNR >72%, mean creatinine) from the clinical heuristic [56]. As part of the workflow of the InfEHR framework, we automatically generate labeling rules from the EHR graph structures that are aggregated to produce prior probabilities [71]. The machine generated prior probabilities outperform the clinical heuristic and are comparatively more performant compared to the machine priors generated for CN-S. Given that the rules are generated from nodes which are in turn extracted from EHR sources, we analyze the composition of the rules in terms of node source (e.g., labs, vitals, medications, or clinical notes) [75]. PO-AKI rules selected for aggregation had higher mean presence of nodes from medications and clinical notes (22%, 39% respectively) compared to aggregate rules for CN-S (8%, 17%) which had more representation from vitals and lab results or from unselected rules for PO-AKI (13%, 31%). Taken together with the decreased false negatives (47.5% vs. 79.2% clinical heuristic) these results suggest that InfEHR can learn conditional relational structures that better identify risk in patients who otherwise had low or no indicated risk per the creatinine based clinical heuristic [21]. The GNN component further reduces this uncertainty to produce better rule-in potentials (3.26 GNN vs 1.31 heuristic) [77]. While the PPV is similar, the higher recall of positive cases at greater NPV (0.927 vs 0.824) shows that InfEHR can successfully estimate the required contingencies needed to make good PO-AKI risk assessment and outperform the clinical heuristic (see Supplementary Materials).

InfEHR produced high and comparatively better rule-in and rule-out potentials in the MOVER dataset (rule-in: 6.34, rule-out: 2.32). We examine other metrics that broadly support improvement to the input distribution of probabilistic labels (consistent with the observed better rule-in and rule-out potentials). We use the beta distribution with parameters set to match the prevalence of the empirical sample to serve as a null model. The clinical heuristic’s close alignment with the beta distribution (0.8129) combined with its low negative log loss (0.0360) but high positive log loss (3.2807) reveals that predictions are being made in a "calibrated conservative" pattern that readily rules out but require more evidence to rule in.

In contrast, the revised InfEHR probabilities diverge from the beta distribution (1.5930) while achieving superior performance, suggesting the model has learned additional signal in the data beyond what is captured by clinical heuristics, enabling more confident predictions in both positive and negative cases while maintaining accuracy. The increasing perplexity values from heuristic (1.0730) to InfEHR’s revised probabilities (1.5327), indicates the model learns to express more varied probabilities across cases. When viewed alongside the improved discriminative metrics, this suggests the higher perplexity reflects better-calibrated probability assignments that more accurately capture case-specific uncertainty while maintaining strong overall performance which broadly supports that InfEHR can learn effective features from EHRs automatically without requiring expert provided labels. The positive log loss also shows marked improvement from the clinical heuristic (3.2807 to 1.0828), also showing enhanced accuracy specifically in positive case prediction, though with a measured increase in negative log loss (0.0360 to 0.2839) which is the objective of InfEHR.

InfEHR yields probability distributions with high rule-in and rule-out potentials, exceeding clinical heuristics in all settings (PO-AKI (MOVER and MSHS) and CN-S), because it approaches the learning task from a generative perspective and supplies a wealth of representational capacity in the patient EHR graph. The GNN architecture in InfEHR can effectively capitalize on this as it finds as features temporal relationships between clinical entities. The initial input EHR graph contains all possible temporal relations between the entities (see Supplementary Materials for detail on entity identification) but the GNN learns to coalesce individual entities into high level semantic groupings connected by condensed edges through a pooling operation (e.g., the GNN may automatically learn that receiving an ACE inhibitor and a finding of a certain blood pressure should be coalesced into an abstract singular clinical concept). The training objective enforces the generative paradigm in which the returned probability of an EHR given a disease latent is determined by the certainty that the disease latent is evidenced by the learned features relative to the empirical estimate of the likelihood of the disease itself as learned during the training process. The likelihoods therefore reflect the model’s certainty in how well-evidenced a given disease is by the underlying EHR, which can be interpreted similarly to the results of a clinical test. By returning likelihoods that explicitly model the probability of the disease itself based on the available evidence in the EHR (and not a decision boundary) InfEHR returns likelihoods with uncertainties that can be used in clinical decision making.

The likelihoods are ultimately derived from quantifying interrelationships between clinical variables over time, which as we suggest in the case of CN-S, can provide a basis for clinical inference in settings lacking specific individual biomarkers, or in the case of PO-AKI where dynamically accumulating risks interact with other conditional risks. While clinical evolution is generally acknowledged to be an important phenotypic and prognostic indicator, the complexities involved in characterizing it limit its practical use beyond a general concept. It is not always known what extent of similarity (or over which components) is needed to distinguish like from unique trajectories. This phenomenon is known as sequence explosion where individual variation leads to large numbers of detected phenotypes with shallow support thereby limiting their comparisons [91]. InfEHR overcomes sequence explosion and brings temporal dynamics into a practical reality through several overlapping mechanisms that cooperate to produce an aggregated view of individual temporal dynamics through which comparisons and inferences can be made.

While we suggest the strength of minimizing human bias by learning graph structures from naive temporal graphs, it comes at a computational cost. The edge count in our graphs explodes with the length of EHR modeled which may impede the training of the pooling and self-attention mechanisms at long durations. While we learn on EHR graphs from hospitalized patients with a maximum of 11-day intervals, it may be required to adapt the framework for longer-term temporal modeling, especially in high measurement frequency environments like the ICU. Future work should be done to decide the optimal method. However, a possibility is to treat the maximum EHR length as a hyperparameter and then aggregate the representations from InfEHR over such periods. Other possibilities include removing edges outside of a given temporal range to reduce graph complexity. Future work will also include code and workflow optimizations to support productionizing InfEHR.

While we have focused on the capacity of InfEHR to produce low entropy likelihoods, additional information could be gained from examining high entropy results which point to uncertain predictions by InfEHR. While these may represent limits to algorithmic performance, they may also express information about the instant case where, for example in PO-AKI, a low entropy prediction for a known positive case may point to an individual who experienced specific adverse events during surgery that resulted in AKI, or in the setting of CN-S, an uncertain prediction for a known positive may indicate that the individual never had CN-S despite being treated for it (See note on label fidelity). This facet suggests an added research-based use for InfEHR alongside our suggestions for clinical decision support. Here we show InfEHR to be a performant and scalable method for extracting insights from EHRs which we make widely available (see Code Availability).

## METHODS

### Overview of InfEHR

The premise of InfEHR is that more information is available than is used in individual clinical decision making. The complexities of obtaining information from EHRs limit their utility. InfEHR is a geometric deep learning approach for resolving clinical uncertainty using EHRs with minimal human intervention. The framework is designed to perform in realistic clinical settings where large volumes of labeled training data cannot be obtained and where existing knowledge is limited.

InfEHR is composed of three modules used sequentially (1) automatic determination (Node Discovery) and embedding of relevant clinical events from EHRs. The nodes are connected according to the naive temporal ordering in the EHR forming EHR graphs. an attention-based graph neural network (GNN) embeds EHR graphs using self-supervision. These embeddings, representing the complete patient record, are used in an automatic rules generation engine to obtain initial likelihoods for all unlabeled cases, uncertainty in these likelihoods is resolved through semi-supervised training of the GNN using a specialized loss function.

We explain each component, providing detailed equations, and provide details of the datasets in the following sections.

### Data

#### Training Dataset

We obtain structured and unstructured data from 11 million electronic health records (EHRs) from the Mount Sinai Health System stored in the Mount Sinai Data Warehouse (CN-S) and through the Extrico platform (PO-AKI) over time varying measurements, medications, and clinical progress notes.

We obtain records for potential neonatal CN-S cases by identifying individuals with at least 48 hours of antibiotic exposure administered in the NICU. We then abstract all antibiotic courses for such individuals meeting this requirement (n=8067, 9256 antibiotics courses). A physician subject matter expert manually confirmed the CN-S status for 3653 antibiotic courses, but 5091 were ultimately used for model training where label information was not required (See unsupervised training as described below).

We obtain records for individuals undergoing surgery of any kind with pre-surgical hospitalization >= 2 days and who had in-hospital serum creatinine measurements taken at 72 hours (n=22,138) post operation to compute AKIN scores. For patients with multiple surgeries, we consider only first surgeries with subsequent operations > 72 hours (n=8031). We assigned a positive AKI diagnosis to any patient with AKIN score > 0.

#### Validation Dataset

We use the EPIC EHR cohort in the MOVER dataset (n= 39,685) and include only patients with serum creatinine measured pre-operatively (2 or more measurements) and and at 72 hours post-operatively (n=2631). We apply the AKIN definitions to obtain labels.

#### Data Preprocessing

We apply pre-processing steps to the CN-S and PO-AKI training datasets (individually) and apply the results from the PO-AKI data to the validation dataset.

EHRs with categorical missingness (e.g., no vitals) were excluded from training where we compute likelihoods (retained cases: n=5091 CN-S, n=4267 PO-AKI) however we use all valid records, including incomplete records, for the density estimations required by the node discovery process (see Method of Node Discovery below).

We include vitals measurements and laboratory results measured on at least 100 unique individuals and with representation from both labels (i.e., the measurement does not by itself identify a case). As a result, given only 137 patients with confirmed CN-S, we consider the vital signs of respiratory rate, spO2, temperature, pulse, and systolic / diastolic blood pressures to avoid bias from measurement type. We retain 25 unique vitals in PO-AKI. We consider 387 and 72 unique labs, 47 and 280 distinct medications in CN-S and PO-AKI respectively.

We further process continuous numerical values by dropping any value exceeding more than three times the maximum or minimum clinical reference range (deemed to be likely artifactual). Non-numerical observations corresponding to lab results were standardized by applying Levenstein distance to coalesce all similar variations to the most frequently observed term. We process categorical features derived from clinical notes as follows: we apply a UMLS matcher to identify terms from clinical notes matching a UMLS term with high confidence. The extracted terms are further filtered in the node discovery process.

### Deep Geometric Learning Approach

#### Notation

We represent individual EHRs as directed graphs by determining relevant clinical events *ɛ* (such as a measurement value within a certain range, or the appearance of a term in a clinical note) where *ɛ* is discovered through an automatic process (Node Discovery).

We derive embeddings for these clinical events by learning a manifold Μ comprised of all events *ɛ* and their respective semantic types *τ*. We apply an operator (here concatenation) to obtain the representations of all possible clinical events as *E* = {*ϕ*(*ɛ* ∈ *M*, *τ* ∈ *M*): ℝ^*n*^ → ℝ^*d*^}.. Graphs representing patients are constructed by identifying the time stamp of *ɛ* ∈ E in the patient record, embedding the time stamp using a network trained on the date2vec objective and concatenating this to *ɛ* resulting in initial node embedding *h*_*i*_^(0)^ ∈ ℝ^*m*^ (*m* > *d* > *n*). The graph is defined as set of nodes and edges *G* = (*ɛ*, *V*(*t*)) with directed edges *e_i,j_* = (*v*_*i*_(*t*) < *v*_*j*_(*t*))

##### Problem definition

Given the graph *G* we train networks to learn whole graph representations in ℝ^*d*^ for (1) self-supervised representations of EHRs, and (2) computing likelihoods over clinical queries. Exact definitions of the loss functions used for training in (1) and (2) are provided below.

### Construction and Definitions of EHR Graphs

InfEHR computes likelihoods through sequential processing of EHRs. We obtain EHRs and then represent them as temporal graphs *G* = (*ɛ*, *V*(*t*)) according to the definitions below:

#### DEFINITION 1: EHR

Given *R_i_*, comprising all medical records for patient *i* occurring over the set of times *T_i_*, we extract the Electronic Health Record (EHR) of patient *i*, denoted as *ɛHR_i_*:

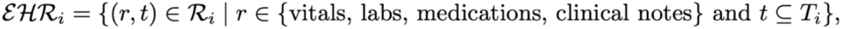

with *t* bounded by:

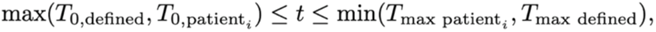

Here, *T*_defined_ indicates the timestamp of a clinical event or user-provided temporal duration, and *T*_patient_, corresponds to an observed timestamp in the records of patient *i.* In the case of multiple defined clinical events occurring within the records of patient *i,* each event results in a unique EHR identified by *ɛHR_ij_*.

#### DEFINITION 2: EHR Graph

We take EHRs (see Definition 1) and represent them as temporal graphs. We discover and collate nodes from the collected EHRs using unsupervised methods into a global node pool (Nodes), embed individual time stamps using Time2Vec (Times), and form temporal edges following:

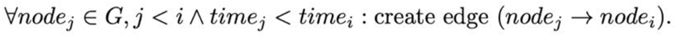

#### Algorithm 1

Construct EHR Graph

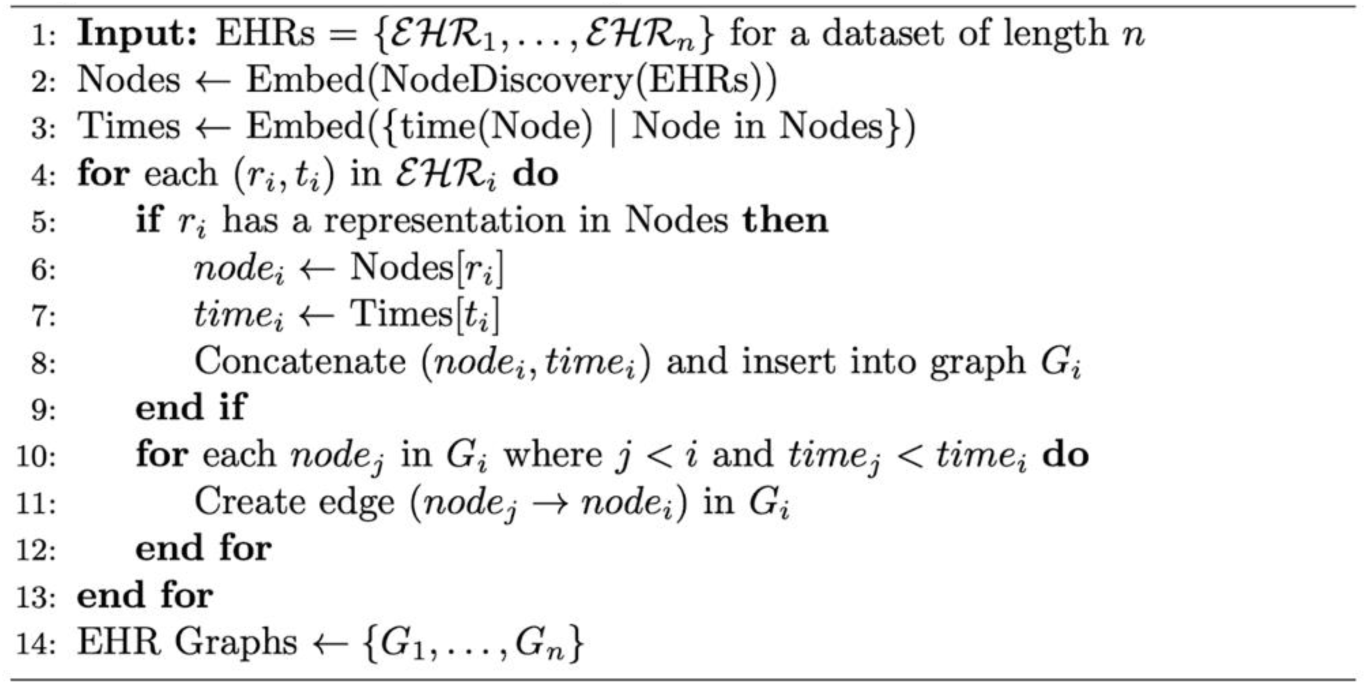

### Method of node discovery

We discover the set of nodes comprising the global pool of clinical events as nodes *E* using density-based selection procedures (Node Discovery). We apply this process to the CN-S and PO-AKI datasets separately. We use the learned results from the training dataset to extract clinical events from the validation dataset.

We define the operations involved as follows:

#### Continuous Variables

We fit kernel density estimations (KDE) to the set of all observations for all EHRs for each measurement type (e.g., heart rate, respiratory rate, white blood cell count etc.). The resulting KDE curve indicates local densities by the intervals between local peaks which we then use to discretize the continuous measurement. The number and distance between peaks is set by a single bandwidth parameter which we determine empirically to satisfy the following constraints: the discretization must be shared by at least 100 unique individuals (e.g., a local density for blood glucose must contain measurements observed for at least 100 unique individuals) while maximizing the number of identified intervals.

#### Discrete Variables

InfEHR uses discrete but time varying information including medications and clinical terms in notes. We include medications that were administered to at least 100 unique patients that are in the prescribable subset of RxNorm. We apply a selection process to terms extracted from clinical progress notes.

To identify potential nodes within these clinical notes we first align all notes with the UMLS ontology by applying QuickUMLs and extracting all terms matching to a UMLS term with high confidence. We weight the collection of extracted terms using TF-IDF, then apply NMF with automatic determination of latent topic number (components in H). We analyze the resulting low rank term-weight matrix to identify and retain terms strongly associated with any latent topic (>= top 10% of distributed topic weights). This procedure simultaneously selects terms based on frequency and informational content.

The overall node selection process automatically compresses the range of all clinical events to a subset based on the underlying density distributions of the instant dataset.

### Method of node representation

We derive 64-dimensional numerical representations (embeddings) for the identified nodes described above. To compute the node embeddings, we first construct a bipartite graph with partitions over individual patients and the identified global set of nodes. Next, we compute the overlap weighted projection of the clinical event nodes over the patients and retain only edges weighted at or above the 25th percentile edge weights. We add nodes representing semantic types (e.g., the name of a lab measurement or vitals sign category) to the projected graph and connect them to relevant nodes with maximum edge weight. The neighborhood of any individual node therefore includes all nodes of the same semantic type as well as nodes across semantic types with high co-occurrence (indicated by high-weight edges). Nodes are encoded to reflect neighborhood information using the node2vec algorithm.

The resulting collection of embeddings (including clinical events and semantic identifiers) forms a manifold which naturally encodes semantic clinical relationships into spatial distances between embeddings. We assign to each node in an EHR graph the resulting relevant embedding, subject to some added components as described below. Note: learn embeddings from the training datasets individually, we use these embeddings in the validation dataset without re-training.

### Tuning the general node representation to individual temporal contexts

After extracting the set of clinical events, we extract the time stamp of their occurrences in individual EHRs. We adjust all time stamps to reflect elapsed times by subtracting the earliest time stamp corresponding to a clinical event in each EHR. We aggregate all unique time stamps for all EHRs and derive 32 dimensional embeddings for each time stamp using the time2vec algorithm:

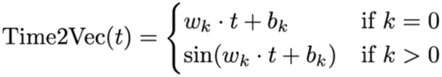

where:

- *t* is the time component (like timestamp, hour of day, etc.)
- *w_k_* and *b_k_* are learnable parameters of the model.

The general representation of a clinical event is formed by concatenating the event embedding with the semantic type embedding (e.g., the embedding of a certain KDE density for blood glucose is concatenated with embedding for blood glucose). These generalized embeddings (consistent across patients) are tuned to the individual by adding the embedding of the time stamp of its occurrence (particular to the patient). The resulting embeddings render clinical events into a machine-readable format. While we do not explicitly use time stamps as positional markers, the vectorization of time adds temporal information into the representation of a clinical event. The numerical distance between locally co-occurring but semantically distinct clinical events is reduced by the similarity of their time stamp components compared to events more separated in time. Individual variation in temporal dynamics therefore shapes the representation of the clinical event to the machine, transforming generalized clinical event representations to reflect individual contexts.

### Training the GNN on attributed EHR Graphs

We train a GNN to produce whole graph embeddings subject to additional processing layers under a self-supervised and semi-supervised objective (details below). We use a consistent architecture adapted to supervised and self-supervised training regimes.

### Self-Supervised Learning for Whole Graph Representations

We learn whole graph representations by training the GNN using a self-supervised loss as described below:

#### SSL Training Algorithm

Detailed Graph. Self-Supervised Learning with VICReg and MI

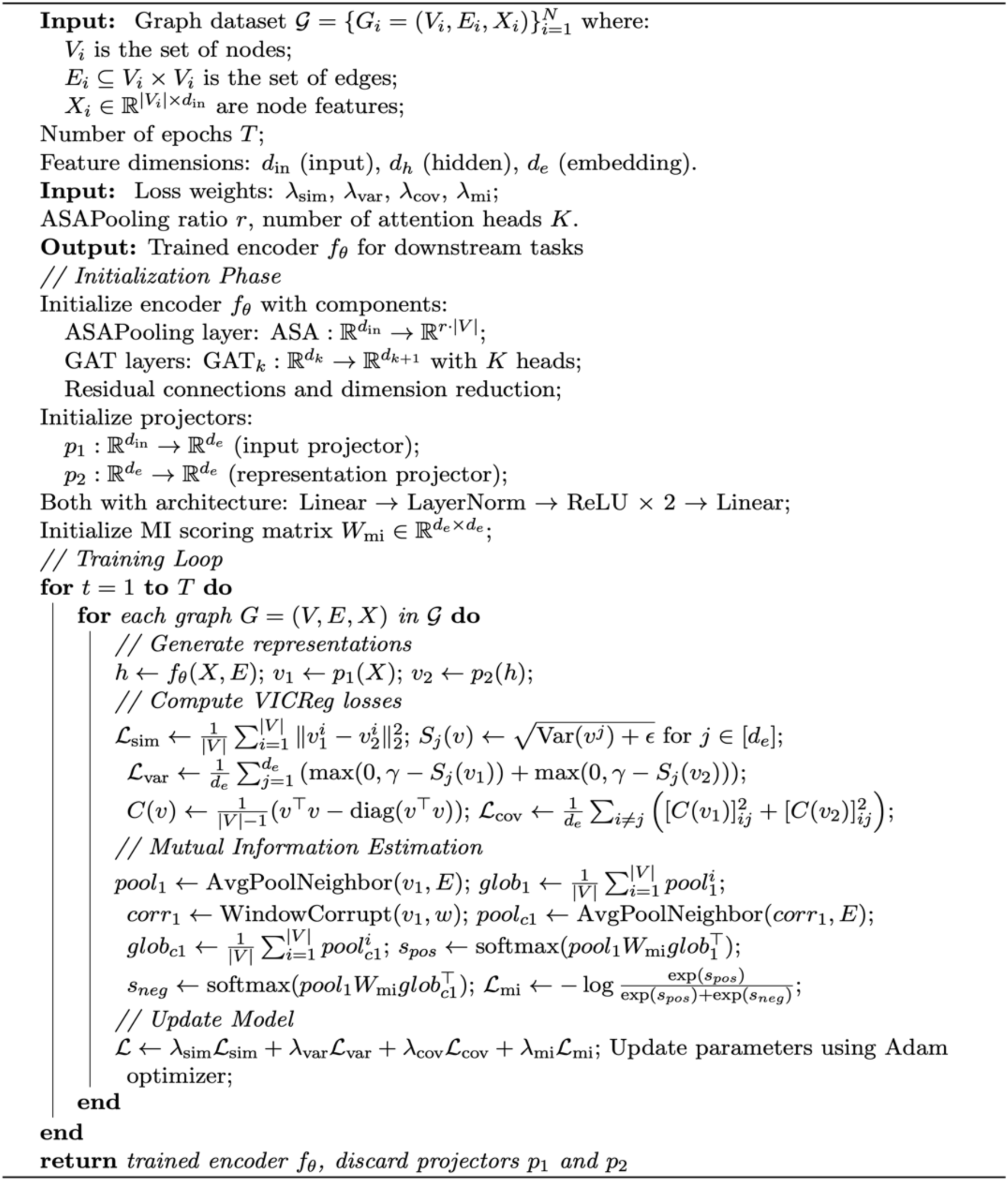

Our SSL training algorithm uses a VICReg framework [85], more commonly used for image encoding, enhanced with mutual information (MI) estimation tailored to graph data. The procedure initializes an encoder with ASAPooling and GAT layers, along with two projectors for input and representation transformation. During training, each graph generates two views: raw features projected through p₁ and encoded features through p₂. The projectors, implemented as multi-layer perceptrons with normalization and non-linearities, serve as learnable transformations that map the input and encoded representations to a shared embedding space while preventing information collapse. This architectural choice enforces an information bottleneck that prevents the encoder from learning trivial solutions, while the projectors’ flexibility allows optimization of the contrastive learning objective without constraining the encoder’s representation capacity. Post-training, the projectors are discarded, preserving the encoder’s learned manifold structure for downstream tasks.

The loss function combines four components: similarity loss (ensuring view alignment), variance loss (preventing dimensional collapse), covariance loss (decorrelating features), and mutual information loss (maximizing node-to-graph information while minimizing it for corrupted samples). The MI estimation uses a structured corruption scheme and InfoNCE-style loss computation.

The corruption scheme generates negative samples by shuffling node features within windows of the EHR graph. This amounts to introducing random and unrealistic relationships and orderings between clinical events. The MI estimation encourages the model to distinguish valid clinical structures and their representations from these unrealistic examples.

The total loss is weighted as L = λsimLsim + λvarLvar + λcovLcov + λmiLmi, optimized using AdamW. After training through T epochs (we trained for 1000 epochs), the encoder is preserved for downstream tasks while projectors are discarded.

This SSL loss function encourages EHR representations that capture local temporal patterns within patient records and also global patient state allowing for encodings that capture inter-patient variation (differing global states) simultaneously with encoding shared local temporal structures. This promotes meaningful semantics in several ways: high-density regions are likely to represent patients with common clinical patterns or disease trajectories, while sparse regions may indicate rare conditions or unique patient presentations. We use the spatial distances to infer the disease state as follows.

### Deriving Instance Level Priors Automatically

#### Label propagation using self-supervised embeddings

We train the GNN encoder to produce self-supervised embeddings as above and apply label propagation as described by Zhuo et al (2003) and implemented in sci-kit learn. We hypothesize that training on a self-supervised objective, as described above, results in automatic alignment of phenotypically similar people such that the assumptions of label smoothing, which is that semantic similarity is a function of spatial distance, can be met.

The label smoothing algorithm iteratively learns a smooth classification function whereby we take the 110 labeled samples and spread label information to spatially proximate samples and label each sample according to the flow of labels it receives during the propagation process. We apply the label spreading algorithm using an RBF kernel (gamma at 70) to determine probabilistic distances between embeddings and set the clamping parameter alpha, controlling the relative importance of the initially labeled examples in deciding the predicted labels for unlabeled examples, to 0.5 based on previous experience. We achieved 0.18 (CN-S) and 0.34 recall (PO-AKI) with precision of 0.67 and 0.78 respectively (outperforming the clinical heuristic in both cases) suggesting the spatial similarity assumptions appreciably obtained in the self-supervised embeddings.

#### Integration of Spatial Information and Structural Information

We derive weak labeling heuristics from structural features of EHR graphs using label information provided from label propagation over the self-supervised embeddings (spatial information as described. Ordinarily such weak heuristics are generated by a human expert which involves bias and challenges in precisely these clinical settings where existing clinical knowledge is limited. We present a method to automatically generate them at scale in uncertain conditions. We find that a random sample of automatically generated heuristics were corroborated by existing literature [85], [86], [87]. Another potential application of InfEHR could be hypothesis generation from predictive weak heuristics.

### Weakly-Supervised Learning Over Uncertain Priors

Given an EHR Graph *G* = (*V, E*), the model initially condenses and rewires the graph through a learned pooling operation resulting in:

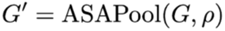

We derive the global representation X and logits for *G′* as follows:

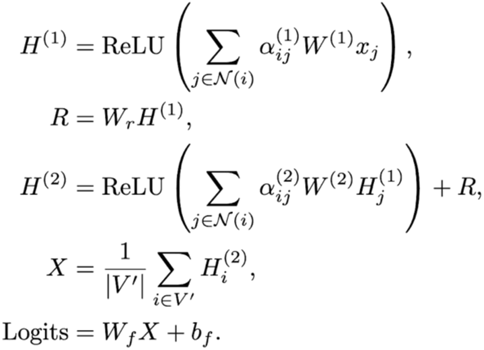

The ASAPooling operation [82] uses an attention-based mechanism to derive cluster medoids and assign cluster memberships for nodes over a fixed receptive field to produce a new, pooled graph. Clusters are scored for inclusion in the pooled graph and re-connected with edge weights that indicate the topology of the original graph. We apply a message passing algorithm over the pooled graph using graph self-attention to compute new node representations successively. The node representations are derived by learning an attentional coefficient that weighs the relative importance of a node to its neighbor in the aggregation phase of the message passing algorithm. To avoid over-smoothing of node representations we apply a residual between successive message passing steps. Finally, to obtain the whole graph representation we take the mean over node features for all nodes in the pooled graph resulting in a single high dimensional vector. We further process the whole graph representation using linear layers to return likelihoods according to the following loss criterion.

### GNN Training with Feature-Based Weighting of Kullback-Leibler Loss

We train the GNN (previously described) under our own loss function similar to the RQ loss proposed by Rolf et al and including a small network to learn example specific loss weighting functions. RQ loss consists of a generative formulation allowing the optimization of the log-likelihood of learned graph features relative to an assumed underlying generative process (here a disease latent). The loss function is consistent with the overall data representation strategy in which we capture disease latents at multiple levels (from initial node encodings to the naive temporal EHR graphs). We extend this function further by learning a dynamic weighting mechanism that continuously adapts during training, learning to adjust sample importance based on evolving patterns in the penultimate layer representations. By modulating the RQ loss through learned weights we increase attention to the most informative samples as the feature space becomes progressively more structured throughout training.

The weighted RQ loss minimizes the following function, jointly optimizing parameters *0* for the primary model and 0 for the weighting network:

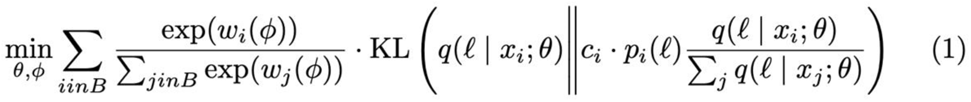

Definitions:

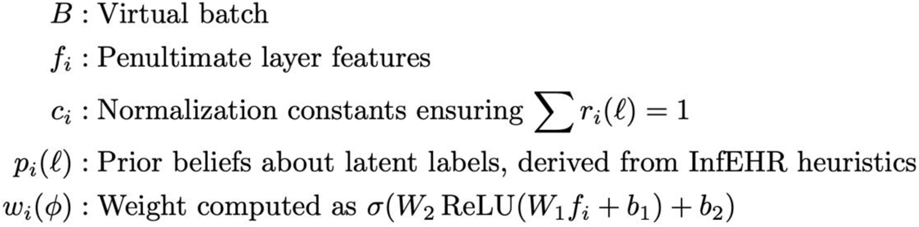

where *ϕ* = {*W*_1_, *W*_2_, *b*_1_*,b_2_}* are trainable weights and biases of the neural network.

Parameters:

- *θ*: Parameters of the primary model estimating *q*(*ℓ* | *x_i_, θ1+)*.
- *ϕ*: Parameters of the neural network calculating *w_i_*.

Optimization by simultaneous updates to *θ* and *ϕ*, aligning the model’s outputs with instance importance in batch *B*.

Notably, the loss function allows us to approximate the complex conditional probability *p*(*x*_*i*_ | *ℓ*|)directly from the empirical sample without the requirements of variational methods which require parameterizing two networks (an encoder and a decoder) or explicit information concerning the link between a feature (here a feature would be a temporal dynamic abstracted from an EHR Graph) and a latent disease.

### Performing Validation

We construct EHRs graphs from the MOVER dataset by first aligning the records in MOVER to the same namespace as the MSHS data (e.g., creating a mapping between the same medication or laboratory measurement with varying names) and then using the learned clinical manifold from MSHS data to extract clinical events from the MOVER records into naive temporal graphs as described previously. Notably, MOVER data does not include clinical progress notes which limits the full translation of MOVER events to the MSHS manifold. All vitals measurement types in the MOVER dataset were in MSHS however some laboratory measurements and medications in MOVER did not have correspondence in MSHS. We omit any such record from the temporal graphs.

To apply the GNN for semi-supervision from MSHS data to graphs from MOVER, we ablate all nodes from clinical text in the MSHS graphs and re-train the GNN on the MSHS graphs. We apply this GNN to the MOVER graphs to obtain initial probabilistic labels (see figure 6, InfEHR priors). We therefore transfer knowledge from previous training in the form of the learned clinical manifold and in the prior probabilities.

**Figure 5:**
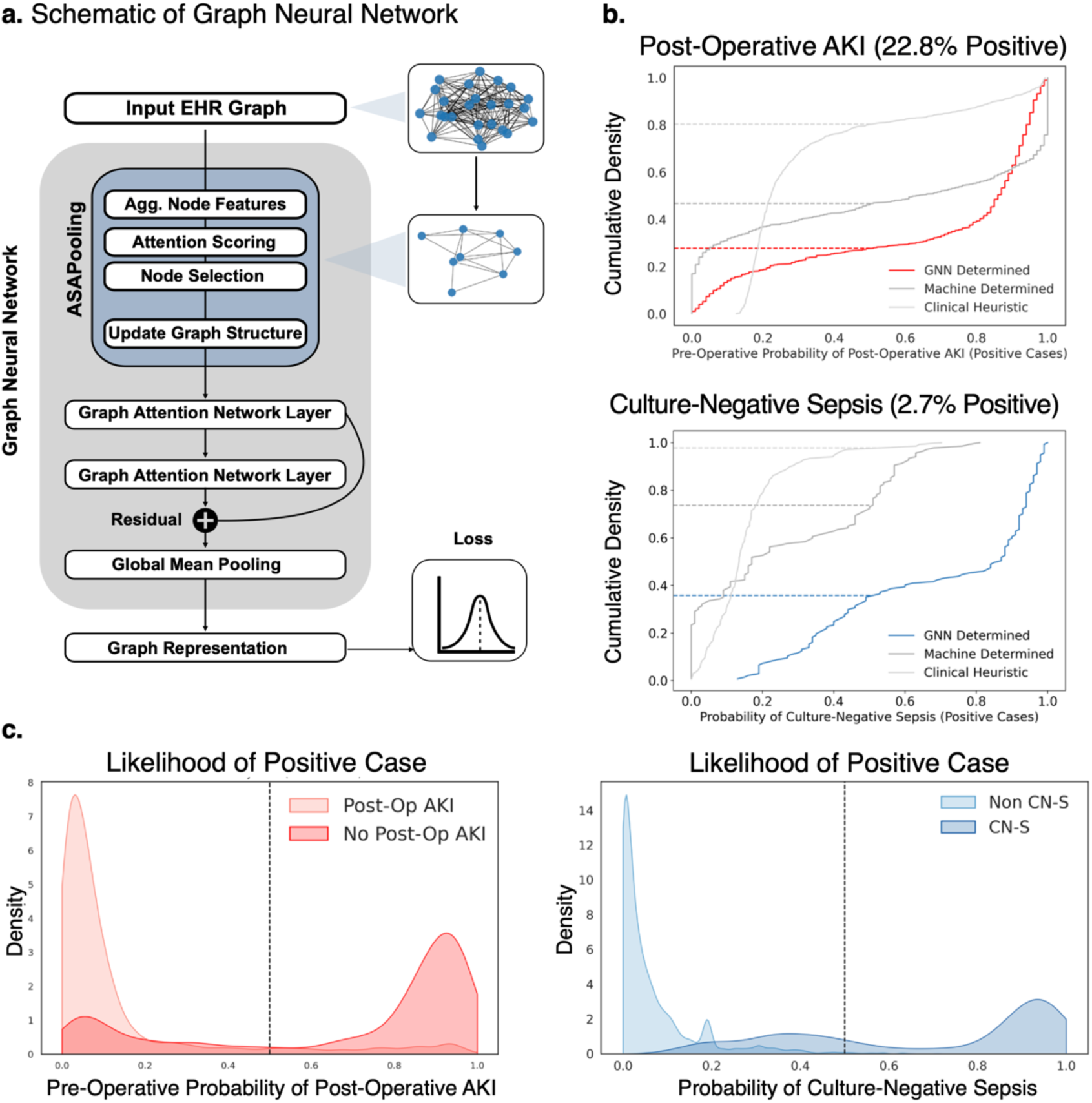
Resolution of clinical uncertainty with InfEHR. **A**. We develop and apply an unsupervised process for the automatic identification of clinical events in EHRs (as nodes) to derive initial EHR graphs. InfEHR learns a new graph structure by coalescing individual nodes (relating to medications, vitals, labs, and notes) into higher order clinical concepts with learned connections. The resulting machine derived graph is then processed with graph attention mechanisms which are trained end to end to relate structural features in the graphs to disease likelihoods **B.** InfEHR significantly improves prior probabilities and outperforms clinical heuristics in identifying positive cases under realistic levels of moderate and severe class imbalances. **C.** While established clinical heuristics readily rule-out conditions, the probability distributions from InfEHR in both disease settings show that InfEHR assigns low or no likelihood of positivity to negative cases while assigning high likelihoods to true positives and are therefore effective for the challenging clinical task of ruling-in a low frequency condition. InfEHR also reproduces its uncertainty in the form of high entropy likelihoods, which may point to important subpopulations for research, and to cases for which InfEHR requires more information or examples in clinical applications.

**Figure 6:**
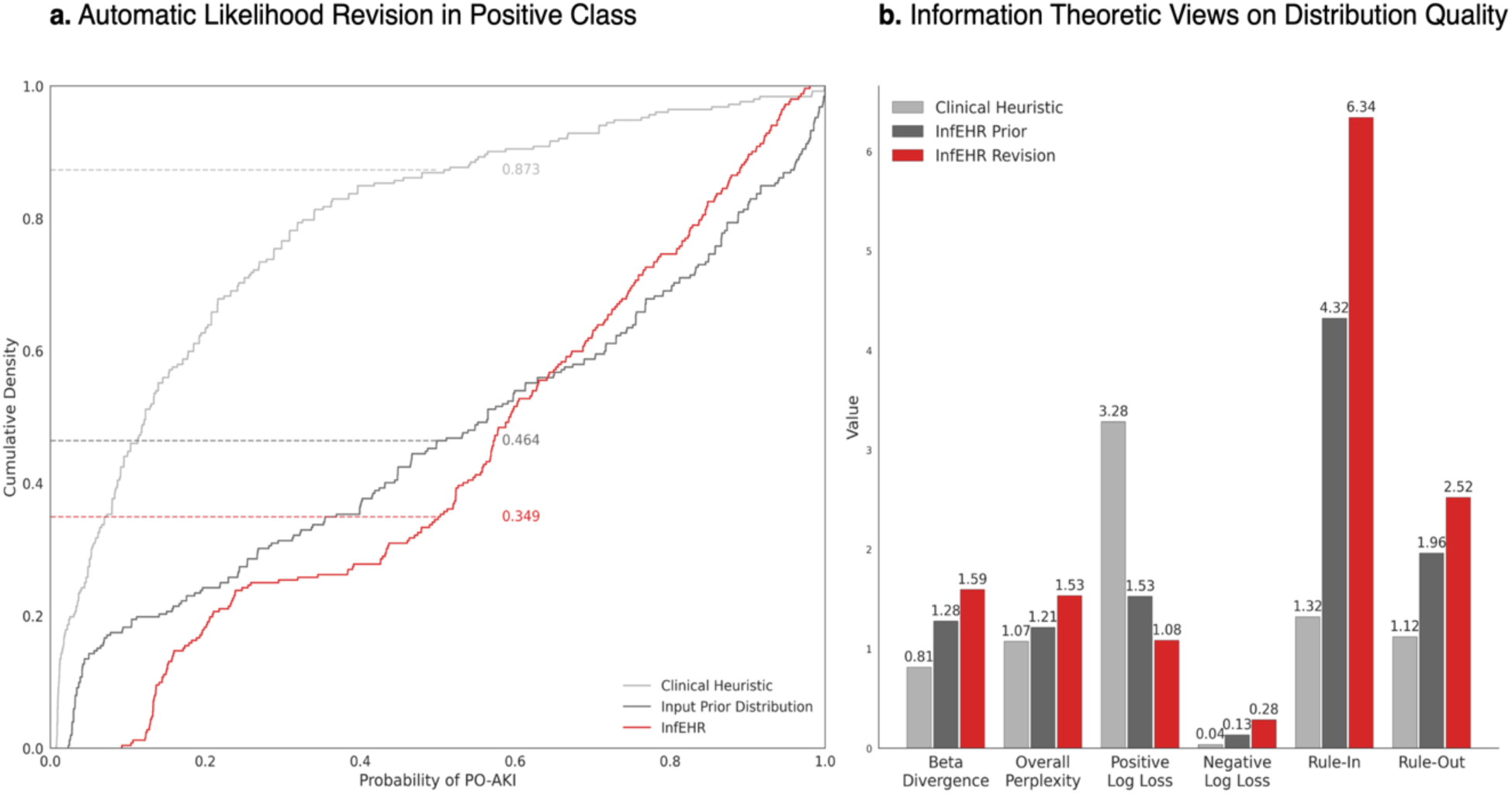
InfEHR revises priors obtained from different domains to local distributions without human intervention. **A**. We evaluate the performance of InfEHR in positive case identification by examining the distribution of likelihoods for positive cases (performance in negatives reported elsewhere). We apply the PO-AKI heuristic with a threshold of 0.5 to a new cohort and obtain similar but slightly worse results (<15% positives identified, in gray). Performance differences reflect underlying clinical and epidemiological differences (e.g., lower disease prevalence) between cohorts. The previously trained GNN is applied to the new and unlabeled cohort to obtain PO-AKI likelihoods (in black). The resulting likelihoods outperform the clinical heuristic leading to identification of 54% of positives. InfEHR further revises these initial likelihoods to identify 66% (in red). We report results using the naive threshold of 0.5 however in practice lower thresholds can be used with InfEHR to identify more positives and maximize clinical net benefit. **B.** The increasing perplexity from heuristic (1.0730) to InfEHR Revision (1.5327) reflects a shift to more varied, case-specific likelihood assignments. While the heuristic’s lower perplexity indicates conservative predictions more aligned with the beta distribution (0.8129), InfEHR’s higher perplexity represents more nuanced probability assignments that better discriminate between cases. This is evidenced by dramatically improved positive log loss (3.2807 to 1.0828) despite a modest increase in negative log loss (0.0360 to 0.2839), suggesting the model learns to express appropriate uncertainty while maintaining strong predictive performance (rule-in: 6.34, rule-out: 2.52). These metrics support that InfEHR can self-improve prior distributions and automatically learn features most relevant to making predictions in local populations.

**Figure 7:**
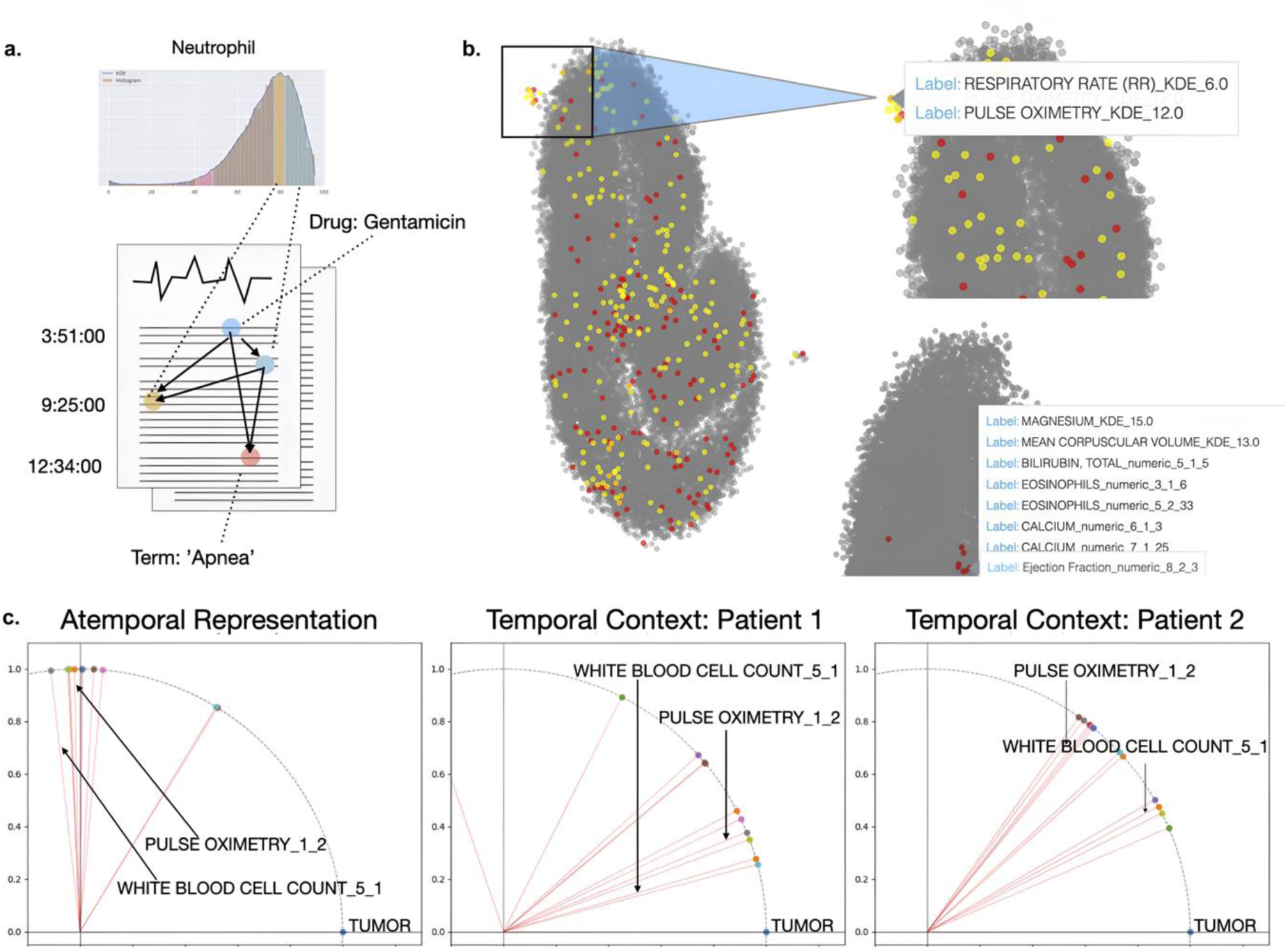
Electronic Health Records (EHRs) are represented automatically as EHR graphs through an unsupervised process. The EHR graph contains a compact record of all clinical events (nodes) and their temporal relationships (edges) over time varying aspects of EHRs including vitals and labs measurements, clinical notes, and medications. We do not assume what nodes may exist but discover them **a.** InfEHR identifies clinical events according to the underlying data. We develop nodes in the graphs as follows: A KDE based heuristic identifies local densities (often automatically recapitulating clinical reference ranges) in continuous measurements as nodes. Terms from clinical notes are identified by alignment with UMLS followed by an NMF based filtering routine. Nodes are aligned to individual records following identification of the node and time stamp in an individual record. Edges are formed between nodes so that each precedent node has a directional edge to every antecedent edge leading to an unbiased but densely connected temporal graph. **b.** Node representations are computed using a mono partite projection over a patient-node bipartite graph. The collection of node representations comprises a semantic manifold which preserves important clinical relationships into a vector encoding (see: respiratory rate and pulse ox.) **c.** We embed time stamps and attach them to node embeddings so that the general representation (see Atemporal Rep.) is tailored to the individual (compare angular distances of the same nodes in Patient 1 and Patient 2). This allows InfEHR to automatically encode temporal dynamics which can be used for learning phenotypic features by the GNN.

InfEHR is designed to learn dynamical temporal features that can be used for clinical uncertainty reduction. To show that InfEHR does this we train a GNN on MOVER graphs (n=2426) with probabilistic prior labels obtained from previous training on MSHS graphs and without any human provided labels. This parallels discriminative model training while being consistent with the InfEHR loss function and framework. Using these priors (without any human provided labels) we train for 20 epochs (by early stopping criterion). We use this GNN to compute final likelihoods (see table and figure 6).

## Data Availability

The clinical data reported in this study cannot be deposited in a public repository because they are confidential medical records however data contained in MOVER is available for download following completion of a data usage agreement. PyTorch implementation together with documentation, computational intermediates, and examples of usage is available in the GitHub repository.

https://mover.ics.uci.edu/index.html

https://github.com/Nadkarni-Lab/InfEHR

## Code Availability

PyTorch implementation together with documentation is available at https://github.com/Nadkarni-Lab/InfEHR. The data from MOVER is available at https://archive.ics.uci.edu/dataset/877/mover:+medical+informatics+operating+room+vitals+and+events+repositor by credentialed access.

## REFERENCES

[1] Sackett, D. L., Rosenberg, W. M., Gray, J. A., Haynes, R. B., & Richardson, W. S. (1996). Evidence based medicine: what it is and what it isn’t. BMJ, 312(7023), 71–72.

[2] Djulbegovic, B., & Guyatt, G. H. (2017). Progress in evidence-based medicine: a quarter century on. The Lancet, 390(10092), 415–423.

[3] Bate, L., Hutchinson, A., Underhill, J., & Maskrey, N. (2012). How clinical decisions are made. British journal of clinical pharmacology, 74(4), 614–620.

[4] Hunink, M. G. M., Weinstein, M. C., Wittenberg, E., Drummond, M. F., Pliskin, J. S., Wong, J. B., & Glasziou, P. P. (2014). Decision making in health and medicine: integrating evidence and values. Cambridge University Press.

[5] Timmermans, S., & Angell, A. (2001). Evidence-based medicine, clinical uncertainty, and learning to doctor. Journal of health and social behavior, 42(4), 342–359.

[6] Maxim, L. D., Niebo, R., & Utell, M. J. (2014). Screening tests: a review with examples. Inhalation toxicology, 26(13), 811–828.

[7] Kumar, A., Roberts, D., Wood, K. E., Light, B., Parrillo, J. E., Sharma, S., … & Cheang, M. (2006). Duration of hypotension before initiation of effective antimicrobial therapy is the critical determinant of survival in human septic shock. Critical care medicine, 34(6), 1589–1596.

[8] Fleisher, L. A., Fleischmann, K. E., Auerbach, A. D., Barnason, S. A., Beckman, J. A., Bozkurt, B., … & Wijeysundera, D. N. (2014). 2014 ACC/AHA guideline on perioperative cardiovascular evaluation and management of patients undergoing noncardiac surgery: executive summary: a report of the American College of Cardiology/American Heart Association Task Force on Practice Guidelines. Circulation, 130(24), 2215–2245.

[9] Bose, S., & Talmor, D. (2018). Who is a high-risk surgical patient?. Current opinion in critical care, 24(6), 547–553.

[10] Adler-Milstein, J., Jha, A. K. (2017). HITECH Act Drove Large Gains In Hospital Electronic Health Record Adoption. Health Affairs, 36(8), 1416–1422.

[11] Blumenthal, D., Tavenner, M. (2010). The "Meaningful Use" Regulation for Electronic Health Records. New England Journal of Medicine, 363(6), 501–504.

[12] Longhurst, C. A., Harrington, R. A., Shah, N. H. (2014). A ’Green Button’ For Using Aggregate Patient Data At The Point Of Care. Health Affairs, 33(7), 1229–1235.

[13] Jensen, P. B., Jensen, L. J., Brunak, S. (2012). Mining electronic health records: towards better research applications and clinical care. Nature Reviews Genetics, 13(6), 395–405.

[14] Hripcsak, G., Albers, D. J. (2013). Next-generation phenotyping of electronic health records. Journal of the American Medical Informatics Association, 20(1), 117–121.

[15] Miotto, R., Li, L., Kidd, B. A., & Dudley, J. T. (2016). Deep patient: an unsupervised representation to predict the future of patients from the electronic health records. Scientific reports, 6(1), 1–10.

[16] Rotmensch, M., Halpern, Y., Tlimat, A., Horng, S., & Sontag, D. (2017). Learning a health knowledge graph from electronic medical records. Scientific reports, 7(1), 1–11.

[17] Zhao, J., Papapetrou, P., Asker, L., & Boström, H. (2017). Learning from heterogeneous temporal data in electronic health records. Journal of biomedical informatics, 65, 105–119.

[18] Choi, E., Bahadori, M. T., Schuetz, A., Stewart, W. F., & Sun, J. (2016). Doctor ai: Predicting clinical events via recurrent neural networks. In Machine learning for healthcare conference (pp. 301–318). PMLR.

[19] Bronstein, M. M., Bruna, J., LeCun, Y., Szlam, A., & Vandergheynst, P. (2017). Geometric deep learning: going beyond euclidean data. IEEE Signal Processing Magazine, 34(4), 18–42.

[20] Gainza, P., Sverrisson, F., Monti, F., Rodolà, E., Boscaini, D., Bronstein, M. M., & Correia, B. E. (2020). Deciphering interaction fingerprints from protein molecular surfaces using geometric deep learning. Nature Methods, 17(2), 184–192.

[21] Choi, E., Xu, Z., Li, Y., Dusenberry, M. W., Flores, G., Xue, Y., & Dai, A. M. (2020). Graph convolutional transformer: Learning the graphical structure of electronic health records. In Proceedings of the AAAI Conference on Artificial Intelligence (Vol. 34, No. 01, pp. 606–613).

[22] Velickovic, P., Cucurull, G., Casanova, A., Romero, A., Lio, P., & Bengio, Y. (2018). Graph attention networks. In International Conference on Learning Representations.

[23] Kingma, D. P., & Welling, M. (2013). Auto-encoding variational bayes. arXiv preprint arXiv:1312.6114.

[24] Rajkomar, A., Oren, E., Chen, K., Dai, A. M., Hajaj, N., Hardt, M., … & Dean, J. (2018). Scalable and accurate deep learning with electronic health records. NPJ Digital Medicine, 1(1), 1–10.

[25] Tomašev, N., Glorot, X., Rae, J. W., Zielinski, M., Askham, H., Saraiva, A., … & Sutherland, C. (2019). A clinically applicable approach to continuous prediction of future acute kidney injury. Nature, 572(7767), 116–119.

[26] Beam, A. L., & Kohane, I. S. (2018). Big data and machine learning in health care. Jama, 319(13), 1317–1318.

[27] Grams, M. E., Sang, Y., Coresh, J., Ballew, S., Matsushita, K., Molnar, M. Z., … & Shlipak, M. G. (2016). Acute Kidney Injury After Major Surgery: A Retrospective Analysis of Veterans Health Administration Data. American Journal of Kidney Diseases, 67(6), 872–880.

[28] Shane, A. L., Sánchez, P. J., & Stoll, B. J. (2017). Neonatal sepsis. The Lancet, 390(10104), 1770–1780.

[29] Wynn, J. L., Wong, H. R., Shanley, T. P., Bizzarro, M. J., Saiman, L., & Polin, R. A. (2014). Time for a neonatal-specific consensus definition for sepsis. Pediatric Critical Care Medicine, 15(6), 523–528.

[30] Japkowicz, N., & Stephen, S. (2002). The class imbalance problem: A systematic study. Intelligent Data Analysis, 6(5), 429–449.

[31] Akobeng, A. K. (2007). Understanding diagnostic tests 2: likelihood ratios, pre-and post-test probabilities and their use in clinical practice. Acta Paediatrica, 96(4), 487–491.

[32] Cantey, J. B., & Baird, S. D. (2017). Ending the Culture of Culture-Negative Sepsis in the Neonatal ICU. Pediatrics, 140(4), e20170044.

[33] Zea-Vera, A., & Ochoa, T. J. (2015). Challenges in the diagnosis and management of neonatal sepsis. Journal of tropical pediatrics, 61(1), 1–13.

[34] Kuppala, V. S., Meinzen-Derr, J., Morrow, A. L., & Schibler, K. R. (2011). Prolonged initial empirical antibiotic treatment is associated with adverse outcomes in premature infants. The Journal of pediatrics, 159(5), 720–725.

[35] Ting, J. Y., Synnes, A., Roberts, A., Deshpandey, A., Dow, K., Yoon, E. W., … & Shah, P. S. (2016). Association between antibiotic use and neonatal mortality and morbidities in very low-birth-weight infants without culture-proven sepsis or necrotizing enterocolitis. JAMA pediatrics, 170(12), 1181–1187.

[36] Cantey, J. B., & Patel, S. J. (2014). Antimicrobial stewardship in the NICU. Infectious Disease Clinics, 28(2), 247–261.

[37] Kuzniewicz, M. W., Puopolo, K. M., Fischer, A., Walsh, E. M., Li, S., Newman, T. B., … & Escobar, G. J. (2017). A quantitative, risk-based approach to the management of neonatal early-onset sepsis. JAMA pediatrics, 171(4), 365–371.

[38] Wynn, J. L., & Polin, R. A. (2018). Progress in the management of neonatal sepsis: the importance of a consensus definition. Pediatric Research, 83(1), 13–15.

[39] Ng, S., & Strunk, T. (2021). Biomarkers for late-onset neonatal sepsis. Current Opinion in Infectious Diseases, 34(5), 485–490.

[40] Gilfillan, M., & Bhandari, V. (2019). Biomarkers for the diagnosis of neonatal sepsis and necrotizing enterocolitis: Clinical practice guidelines. Early human development, 138, 104874.

[41] Sharma, D., Farahbakhsh, N., Shastri, S., & Sharma, P. (2018). Biomarkers for diagnosis of neonatal sepsis: a literature review. The Journal of Maternal-Fetal & Neonatal Medicine, 31(12), 1646–1659.

[42] Hofer, N., Zacharias, E., Müller, W., & Resch, B. (2012). An update on the use of C-reactive protein in early-onset neonatal sepsis: current insights and new tasks. Neonatology, 102(1), 25–36.

[43] Chiesa, C., Natale, F., Pascone, R., Osborn, J. F., Pacifico, L., Bonci, E., & De Curtis, M. (2011). C reactive protein and procalcitonin: reference intervals for preterm and term newborns during the early neonatal period. Clinica Chimica Acta, 412(11-12), 1053–1059.

[44] Benitz, W. E. (2010). Adjunct laboratory tests in the diagnosis of early-onset neonatal sepsis. Clinics in perinatology, 37(2), 421–438.

[45] Kheterpal, S., Tremper, K. K., Heung, M., Rosenberg, A. L., Englesbe, M., Shanks, A. M., & Campbell, D. A. (2009). Development and validation of an acute kidney injury risk index for patients undergoing general surgery: results from a national data set. Anesthesiology, 110(3), 505–515.

[46] Hodgson, L. E., Dimitrov, B. D., Roderick, P. J., Venn, R., & Forni, L. G. (2017). Predicting AKI in emergency admissions: an external validation study of the acute kidney injury prediction score (APS). BMJ open, 7(3), e013511.

[47] Thiele, R. H., et al. (2015). Standardization of care: impact of an enhanced recovery protocol on length of stay, complications, and direct costs after colorectal surgery. Journal of the American College of Surgeons, 220(4), 430–443.

[48] O’Neal, J. B., et al. (2016). Acute kidney injury following cardiac surgery: current understanding and future directions. Critical care, 20(1), 187.

[49] Thakar, C. V., et al. (2005). A clinical score to predict acute renal failure after cardiac surgery. Journal of the American Society of Nephrology, 16(1), 162–168.

[50] Park, S., et al. (2015). Impact of electronic acute kidney injury (AKI) alerts with automated nephrologist consultation on detection and severity of AKI: a quality improvement study. American Journal of Kidney Diseases, 65(1), 9–16.

[51] Kellum, J. A., et al. (2021). Acute kidney injury. Nature reviews Disease primers, 7(1), 1–17.

[52] Matheny, M. E., et al. (2010). Development of inpatient risk stratification models of acute kidney injury for use in electronic health records. Medical decision making, 30(6), 639–650.

[53] Nishimoto M, Murashima M, Kokubu M, et al. External Validation of a Prediction Model for Acute Kidney Injury Following Noncardiac Surgery. JAMA Netw Open. 2021;4(10):e2127362. doi:10.1001/jamanetworkopen.2021.27362

[54] Park S, Cho H, Park S, Lee S, Kim K, Yoon HJ, Park J, Choi Y, Lee S, Kim JH, Kim S, Chin HJ, Kim DK, Joo KW, Kim YS, Lee H. Simple Postoperative AKI Risk (SPARK) Classification before Noncardiac Surgery: A Prediction Index Development Study with External Validation. J Am Soc Nephrol. 2019 Jan;30(1):170–181. doi: 10.1681/ASN.2018070757. Epub 2018 Dec 18. PMID: 30563915; PMCID: PMC6317608.

[55] Hodgson, L. E., Roderick, P. J., & Forni, L. G. (2018). Risk prediction models for acute kidney injury in the general hospital population: a systematic review. BMJ Open, 8(4), e022200. doi:10.1136/bmjopen-2018-022200

[56] Hostetter, T. H., et al. (2016). Laboratory values as predictors of acute kidney injury in critically ill patients: a cautionary tale. Nephrology Dialysis Transplantation, 31(10), 1692–1700.

[57] James, M. T., et al. (2021). Derivation and external validation of prediction models for advanced chronic kidney disease following acute kidney injury. JAMA, 326(11), 1074–1087.

[58] Moledina, D. G., & Parikh, C. R. (2018). Phenotyping of acute kidney injury: beyond serum creatinine. Seminars in nephrology, 38(1), 3–11.

[59] Bornstein, B. H., & Emler, A. C. (2001). Rationality in medical decision making: a review of the literature on doctors’ decision-making biases. Journal of evaluation in clinical practice, 7(2), 97–107.

[60] Marewski, J. N., & Gigerenzer, G. (2012). Heuristic decision making in medicine. Dialogues in clinical neuroscience, 14(1), 77–89.

[61] Norman, G. R., & Eva, K. W. (2010). Diagnostic error and clinical reasoning. Medical education, 44(1), 94–100.

[62] Lee WC. Selecting diagnostic tests for ruling out or ruling in disease: the use of the Kullback-Leibler distance. Int J Epidemiol. 1999 Jun;28(3):521–5. doi: 10.1093/ije/28.3.521. PMID: 10405859.

[63] Biggerstaff BJ. Comparing diagnostic tests: a simple graphic using likelihood ratios. Stat Med. 2000 Mar 15;19(5):649–63. doi: 10.1002/(sici)1097-0258(20000315)19:5<649::aid-sim371>3.0.co;2-h. PMID: 10700737.

[64] Grimes DA, Schulz KF. Refining clinical diagnosis with likelihood ratios. Lancet. 2005 Apr 23-29;365(9469):1500–5. doi: 10.1016/S0140-6736(05)66422-7. PMID: 15850636.

[65] Swets JA. Measuring the accuracy of diagnostic systems. Science 1988; 240:1285–93.

[66] Pepe MS. The statistical evaluation of medical tests for classification and prediction. Oxford University Press; 2003.

[67] Klingenberg C, Kornelisse RF, Buonocore G, Maier RF, Stocker M. Culture-Negative Early-Onset Neonatal Sepsis — At the Crossroad Between Efficient Sepsis Care and Antimicrobial Stewardship. Frontiers in Pediatrics. 2018;6:285. doi:10.3389/fped.2018.00285

[68] Bihorac A, Brennan M, Ozrazgat-Baslanti T, et al. National surgical quality improvement program underestimates the risk associated with mild and moderate postoperative acute kidney injury. Crit Care Med. 2013;41(11):2570–2583. doi:10.1097/CCM.0b013e31829860fc

[69] Pencina MJ, D’Agostino RB Sr, D’Agostino RB Jr, Vasan RS. Evaluating the added predictive ability of a new marker: from area under the ROC curve to reclassification and beyond. Stat Med. 2008;27(2):157–172. doi:10.1002/sim.2929

[70] Zhou, D., Bousquet, O., Lal, T. N., Weston, J., & Schölkopf, B. (2004). Learning with local and global consistency. Advances in neural information processing systems, 16(16), 321–328.

[71] Ratner, A., Bach, S. H., Ehrenberg, H., Fries, J., Wu, S., & Ré, C. (2020). Snorkel: Rapid training data creation with weak supervision. The VLDB Journal, 29(2), 709–730.

[72] Rolf E, Malkin N, Graikos A, Jojic A, Robinson C, Jojic N. Resolving label uncertainty with implicit posterior models. In: Proceedings of the Thirty-Eighth Conference on Uncertainty in Artificial Intelligence. PMLR; 2022:1707–1717.

[73] Ioannidis JPA, Bossuyt PMM. Waste, Leaks, and Failures in the Biomarker-Diagnostic Pipeline. Clin Chem. 2017;63(5):963–972.

[74] Hippisley-Cox J, Coupland C. Predicting risk of emergency admission to hospital using primary care data: derivation and validation of QAdmissions score. BMJ Open. 2013;3(8):e003482.

[75] Goldstein BA, Navar AM, Pencina MJ, Ioannidis JP. Opportunities and challenges in developing risk prediction models with electronic health records data: a systematic review. Journal of the American Medical Informatics Association. 2017;24(1):198–208.

[76] Jensen AB, Moseley PL, Oprea TI, et al. Temporal disease trajectories condensed from population-wide registry data covering 6.2 million patients. Nature communications. 2014;5(1):1–10.

[77] Kipf TN, Welling M. Semi-Supervised Classification with Graph Convolutional Networks. arXiv:1609.02907 [cs.LG]. 2016.

[78] Druzdzel MJ, Díez FJ. Combining knowledge from different sources in causal probabilistic models. Journal of Machine Learning Research. 2003;4(Jul):295–316.

[79] Rajkomar A, Dean J, Kohane I. Machine learning in medicine. New England Journal of Medicine. 2019;380(14):1347–1358.

[80] Bossuyt PM, Reitsma JB, Bruns DE, et al. STARD 2015: an updated list of essential items for reporting diagnostic accuracy studies. BMJ. 2015;351:h5527.

[81] Sutton RT, Pincock D, Baumgart DC, Sadowski DC, Fedorak RN, Kroeker KI. An overview of clinical decision support systems: benefits, risks, and strategies for success. NPJ digital medicine. 2020;3(1):1–10.

[82] Ranjan, E., Sanyal, S., & Talukdar, P. P. (2020). ASAP: Adaptive Structure Aware Pooling for Learning Hierarchical Graph Representations. arXiv preprint arXiv:1911.07979

[83] Cantey JB, Pyle AK, Wozniak PS, Hynan LS, Sánchez PJ. Early Antibiotic Exposure and Adverse Outcomes in Preterm, Very Low Birth Weight Infants. J Pediatr. 2018 Dec;203:62–67. doi: 10.1016/j.jpeds.2018.07.036. Epub 2018 Aug 29. PMID: 30172430.

[84] Lopes JA, Jorge S. The RIFLE and AKIN classifications for acute kidney injury: a critical and comprehensive review. Clin Kidney J. 2013 Feb;6(1):8–14. doi: 10.1093/ckj/sfs160. Epub 2012 Jan 1. PMID: 27818745; PMCID: PMC5094385.

[85] Bardes, A., Ponce, J., LeCun, Y. (2022). VICReg: Variance-Invariance-Covariance Regularization for Self-Supervised Learning. arXiv preprint arXiv:2105.04906. Available at: https://arxiv.org/abs/2105.04906

[86] Nelson, C.A., Butte, A.J. & Baranzini, S.E. Integrating biomedical research and electronic health records to create knowledge-based biologically meaningful machine-readable embeddings. Nat Commun 10, 3045 (2019). 10.1038/s41467-019-11069-0

[87] Tingyi Wanyan, Hossein Honarvar, Ariful Azad, Ying Ding, Benjamin S. Glicksberg; Deep Learning with Heterogeneous Graph Embeddings for Mortality Prediction from Electronic Health Records. Data Intelligence 2021; 3 (3): 329–339. doi: 10.1162/dint_a_00097

[88] Nelson, C.A., Butte, A.J. & Baranzini, S.E. Integrating biomedical research and electronic health records to create knowledge-based biologically meaningful machine-readable embeddings. Nat Commun 10, 3045 (2019). 10.1038/s41467-019-11069-0

[89] A foundation model for clinician-centered drug repurposing Kexin Huang, Payal Chandak, Qianwen Wang, Shreyas Havaldar, Akhil Vaid, Jure Leskovec, Girish Nadkarni, Benjamin S. Glicksberg, Nils Gehlenborg, Marinka Zitnik medRxiv 2023.03.19.23287458; doi: 10.1101/2023.03.19.23287458

[90] Salas M, Hofman A, Stricker BH. Confounding by indication: an example of variation in the use of epidemiologic terminology. Am J Epidemiol. 1999 Jun 1;149(11):981–3. doi: 10.1093/oxfordjournals.aje.a009758. PMID: 10355372.

[91] Liu, C., Zhang, K., Xiong, H., Jiang, G., & Yang, Q. (2014, August). Temporal skeletonization on sequential data: patterns, categorization, and visualization. In Proceedings of the 20th ACM SIGKDD international conference on Knowledge discovery and data mining (pp. 1336–1345).

[92] Samad M, Angel M, Rinehart J, Kanomata Y, Baldi P, Cannesson M. Medical Informatics Operating Room Vitals and Events Repository (MOVER): a public-access operating room database. JAMIA Open. 2023 Oct 17;6(4):ooad084. doi: 10.1093/jamiaopen/ooad084. PMID: 37860605; PMCID: PMC10582520

[93] Johnson, J.M., Khoshgoftaar, T.M. Survey on deep learning with class imbalance. J Big Data 6, 27 (2019).

[94] Yaniv Ovadia, Emily Fertig, Jie Ren, Zachary Nado, D. Sculley, Sebastian Nowozin, Joshua V. Dillon, Balaji Lakshminarayanan, and Jasper Snoek. 2019. Can you trust your model’s uncertainty? evaluating predictive uncertainty under dataset shift. Proceedings of the 33rd International Conference on Neural Information Processing Systems. Curran Associates Inc., Red Hook, NY, USA, Article 1254, 14003–14014.

[95] Chuan Guo, Geoff Pleiss, Yu Sun, and Kilian Q. Weinberger. 2017. On calibration of modern neural networks. In Proceedings of the 34th International Conference on Machine Learning - Volume 70 (ICML’17). JMLR.org, 1321–1330.

[96] Hüllermeier, Eyke and Willem Waegeman. “Aleatoric and epistemic uncertainty in machine learning: an introduction to concepts and methods.” Machine Learning 110 (2019): 457–506.

