## Supplemental Figures for "InfEHR: Resolving Clinical Uncertainty through Deep Geometric Learning on Electronic Health Records"

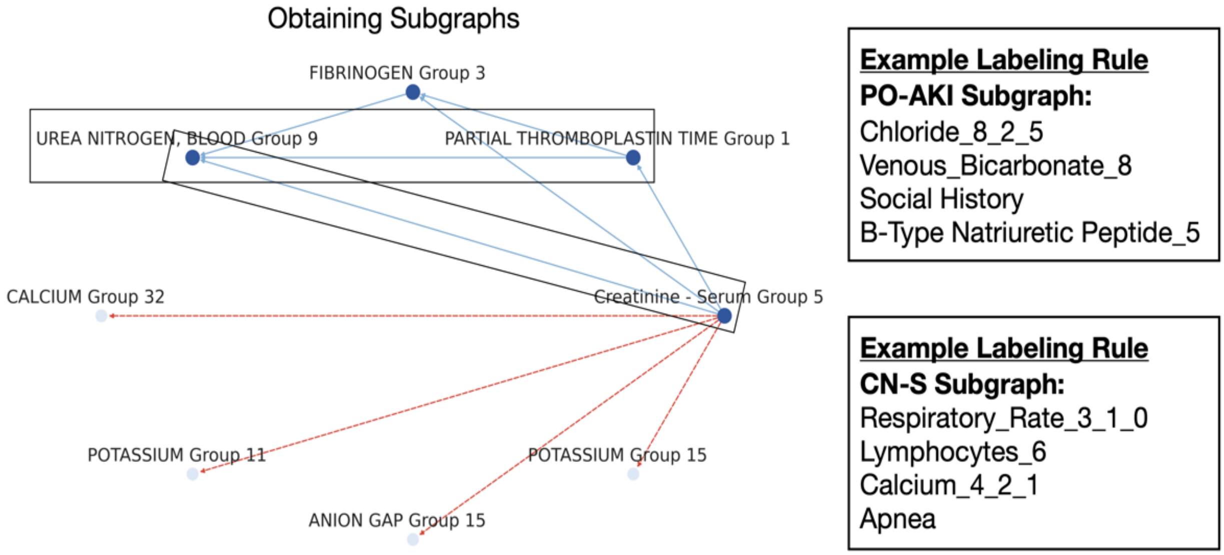


**Supplemental Figure: The graph structure provides a basis to obtain weakly predictive labeling rules in the form of subgraphs.** Initially EHR graphs are embedded following a self-supervised training objective. This results in a coordinate mapping where semantically similar graphs are distributed into spatial neighborhoods. We perform label propagation on the spatial mapping using 110 labeled examples in order to label the entire dataset. We build on these initial labels by selecting subgraphs from labeled examples. Nodes are selected from a 1-hop radius of the initial node. If both the selected node and any number of its near neighbors are disproportionately frequent relative to the selected class the nodes comprise a set of pairwise subgraphs that can be used for labeling.This process selects nodes that co-occur in clinical contexts that occur in the data allowing for useful subgraphs to rapidly be identified **a.** The figure above depicts Serum Creatinine (group 5) and its 1-hop neighborhood (in blue), in red are nodes disproportionately frequent in the opposite class. **b.** Examples of subgraphs (any pairwise combination) obtained through this method for both datatasets.

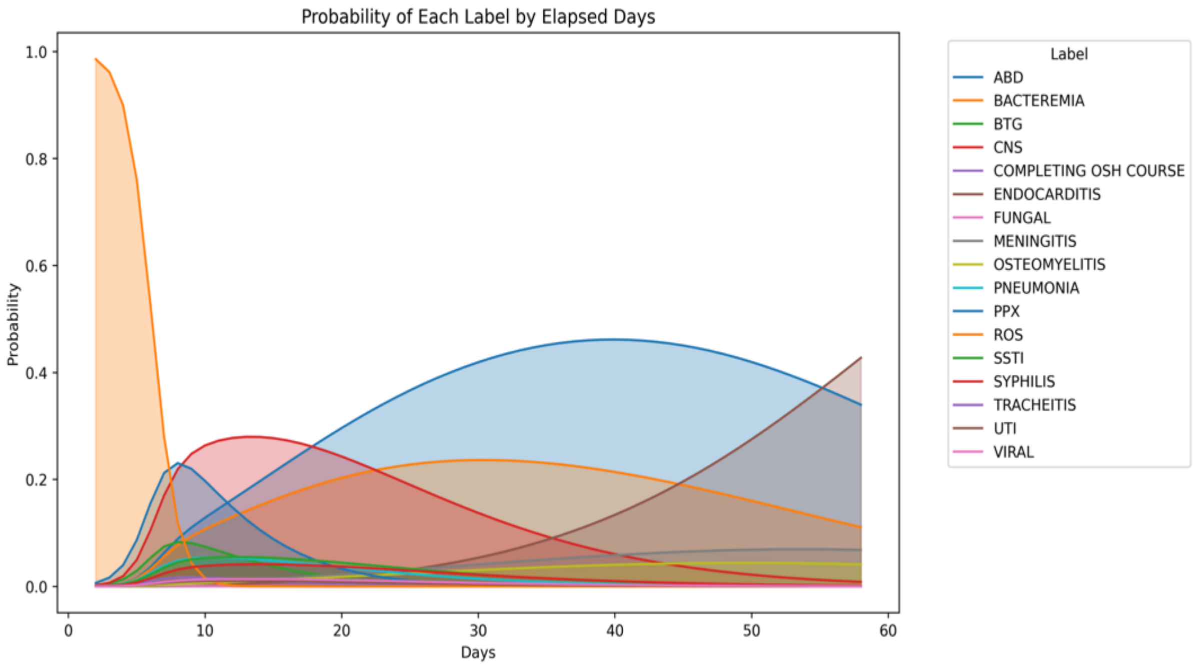


**Supplemental Figure: Uncertainty in identifying culture negative sepsis over time.** We use multinomial logistic regression to estimate and plot the probability of all labels given only the elapsed time with continuing symptoms. In sepsis rule out (ROS) and culture-negative sepsis (CNS) confirmatory positive culture results are never available whereas the mean elapsed time of first positive culture result was 3.28 days for all other case types. We truncate EHR windows to 11 days to preserve uncertainty over all labels and demonstrate performance under clinically realistic settings where the persistence of symptoms may be related to differing underlying causes.
